# Regional brain aging patterns reveal disease-specific pathways of neurodegeneration

**DOI:** 10.1101/2025.11.12.25339989

**Authors:** Pál Vakli, Béla Weiss, Attila Keresztes, Petra Hermann, Alzheimer’s Disease Neuroimaging Initiative (ADNI), James H. Cole, Zoltán Vidnyánszky

**Author notes:** Corresponding authors: Pál Vakli, Zoltán Vidnyánszky. Data used in preparation of this article were obtained from the Alzheimer’s Disease Neuroimaging Initiative (ADNI) database (adni.loni.usc.edu). As such, the investigators within the ADNI contributed to the design and implementation of ADNI and/or provided data but did not participate in analysis or writing of this report. A complete listing of ADNI investigators can be found at: http://adni.loni.usc.edu/wp-content/uploads/how_to_apply/ADNI_Acknowledgement_List.pdf.

## Abstract

The heterogeneity of brain aging is a hallmark of neurological and psychiatric disorders, yet machine-learning tools used to characterize this process, including the ‘brain age’ paradigm, have largely relied on global metrics that lack the specificity to map these complex patterns. Here, we introduce BrainAgeMap, an interpretable deep learning framework that generates fine-grained, voxel-wise maps of brain-predicted age difference (brain-PAD) from T1-weighted magnetic resonance imaging scans. We provide converging lines of evidence for the framework’s clinical, prognostic, and neurobiological utility. Disorder-specific topographies of accelerated aging were identified in Alzheimer’s disease (AD), frontotemporal dementia, and schizophrenia. Longitudinal analysis of the hippocampus revealed accelerated aging in individuals with progressive versus stable mild cognitive impairment (MCI), demonstrating prognostic value. Regional brain-PAD in the temporal lobe correlated strongly with *in vivo* tau pathology measured by positron emission tomography in AD, linking the maps to underlying molecular pathology. Furthermore, regional brain aging in MCI and AD was linked to individual differences in episodic memory function. BrainAgeMap provides a robust tool to delineate disease-specific pathways of neurodegeneration, offering new opportunities for early diagnosis, patient stratification, and monitoring therapeutic interventions.

## 1. Introduction

Brain age prediction has received considerable attention in recent years as a promising method to quantify age-related changes in brain and body health. In this approach, machine learning models are trained to predict the age of healthy individuals based on neuroimaging data^1–4^. The application of the trained models to novel samples revealed that the discrepancy between the predicted and the true (chronological) age—the brain-predicted age difference (brain-PAD)—is increased in certain neurodegenerative and neuropsychiatric disease states and related to various measures of physiological and cognitive function^1–3,5^. Besides its potential clinical utility as a diagnostic aid^6^, as a tool for monitoring disease progression^7,8^ the effectiveness of intervention strategies^9^, brain age prediction may allow a more thorough understanding of the structural and functional alterations that accompany the brain aging process in health and disease.

Most brain age prediction studies rely on T1-weighted brain scans from structural magnetic resonance imaging (MRI) and traditional machine learning algorithms—such as relevance vector regression^10^ and Gaussian process regression^11^—or deep learning architectures, particularly convolutional neural networks (CNNs)^12–14^. A common feature of these approaches is the estimation of a single scalar value, the (global) brain-predicted age. Nevertheless, the structural alterations that accompany brain aging show substantial variability in terms of spatial distribution and rate of progression over time. A recent analysis of more than 100,000 human participants aged between 115 days post-conception and 100 years showed that there was considerable variation across the 32 cortical regions examined, with primary sensory regions peaking earlier and showing faster post-peak declines than frontotemporal association areas.^15^ Large-scale cross sectional analyses also confirmed remarkable regional heterogeneity in cortical thickness trajectories^16–18^, and relatively high interindividual variability in frontal and temporal lobe regions across the lifespan^18^. White matter volume shows continued growth throughout early adulthood with a peak around midlife^15,19,20^, with a subsequent decline that is more pronounced in frontal lobe areas than in posterior regions^21–23^. Besides the spatio-temporal heterogeneity of the normal brain aging process, age-related neurodegenerative disorders—such as Alzheimer’s disease (AD), frontotemporal dementia, etc.—also show, to varying degrees, patterns of brain atrophy that can be detected in T1-weighted MRI scans^24^. Moreover, structural MRI also reveals distinctions as well as commonalities in the neuroanatomical alterations associated with major psychiatric disorders including schizophrenia, bipolar disorder, and major depressive disorder.^25^

Due to the multifaceted nature of brain aging and associated disease states, it is unclear what aspects of this complex process global brain-predicted age reflects, and to what extent. Experimental results show that the neural substrate underlying age predictions cannot be consistently restricted to a few brain regions when the training set covers the entire lifespan^26^, suggesting that there are potentially multiple solutions for brain age prediction from volumetric MRI data^2^. Explainability in brain age prediction is challenging: the interpretation of model parameters in terms of brain processes can lead to erroneous conclusions because these parameters also depend on noise components in the data^27^. Given the apparent complexity of brain aging and the ambiguity in interpreting global brain-predicted age, recent studies introduced models explicitly trained to predict chronological age from brain structure at the local level. Through the generation of individual brain age maps, such local or regional brain age prediction methods may allow for a more quantitative (compared to saliency maps) and fine-grained characterization of age-related neuroanatomical changes in the healthy and diseased brain. Initial work in this field involved brain age prediction at the level of 3D patches^28,29^ or slices^30,31^ However, these methods either provided low spatial resolution^28,30,31^, or the patch-level predictions were aggregated to arrive at a global prediction, without presenting patch-level results^29^. Other approaches included training separate models for each brain region^32–34^. These model ensembles allow for the identification of more detailed structural changes in the aging brain^34^, although they do not incorporate contextual information from neighboring regions and are limited by the specific preprocessing and anatomical atlas used. More recent approaches to local brain age prediction incorporated the use of deep neural networks called U-nets that are typically used for image segmentation^35^. These models allow for the generation of voxel-wise brain age maps without requiring elaborate feature engineering^36–38^. In the first such study, voxel-wise grey and white matter volume maps were divided into 3D blocks, and a U-net was trained to predict voxel-level brain age for each block^37^. Using this method, the authors found evidence for different local brain age patterns in healthy controls and people with mild cognitive impairment (MCI) or dementia. Although the model was capable of generating voxel-level predictions, the effective resolution was lower than a single voxel due to high within-block homogeneity^37^. Subsequent studies employing U-nets also found evidence for differences in regional brain aging in healthy aging and various neurological and neuropsychiatric disease states^36,38^, although they did not perform comparative analyses at the voxel level.

In the current study, we present a novel deep learning-based approach for local and global brain age prediction. This method is based on previous work using the combination of a fully convolutional network (FCN) and a multi-layer perceptron for interpretable Alzheimer’s disease classification^39^. We developed a variant of this framework adapted for brain age prediction, referred to as BrainAgeMap. The FCN in this framework allows for the contiguous evaluation of brain morphology without enforcing global structure onto local predictions^39^, enabling the generation of fine-grained individual brain age maps. We performed stringent comparisons of these maps between healthy controls and people with mild cognitive impairment, Alzheimer’s disease, frontotemporal dementia, and schizophrenia to delineate in detail the putative disease-specific patterns in local brain aging. In addition, we also investigated whether local brain age can be used as a marker of neurodegenerative disease progression. Besides aiding a more mechanistic understanding, longitudinal analyses of brain age have the potential to assess the efficacy of intervention strategies aimed at mitigating the adverse effects of age-related health problems^2^. While the available evidence suggests accelerated global brain aging in Alzheimer’s disease^40,41^ and type 2 diabetes mellitus in older adults^42^, less is known about longitudinal changes in local brain age. Here we focused on the aging of the hippocampus, a pivotal structure for memory formation and consolidation that exhibits marked vulnerability in aging^43,44^.

Besides evaluating local brain age as a putative indicator of disease type and severity, we also investigated its relationship to other imaging and cognitive markers of neurodegeneration. Assessing the relationship between brain-PAD and other physiological and cognitive measures is key to the validation of brain age as an aging biomarker^2^ yet to be examined with local brain age. We focused on the accumulation of tau proteins as measured by positron emission tomography (PET) in cognitively normal (CN) persons and in individuals with MCI or AD. The spatiotemporal pattern of pathological tau accumulation across brain regions has been postulated to be characteristic of different neurodegenerative disorders known as tauopathies, of which AD is the most common^45^. Tau-PET imaging, using the [18F]flortaucipir (FTP) tracer, has shown high accuracy in differentiating AD from other tauopathies^46^ and can be used to predict cognitive performance^47^ and subsequent brain atrophy^48^ in AD. A recent study has shown that global brain-PAD derived from structural MRI was significantly correlated with tau uptake in the temporal lobe in MCI and AD patients^49^. However, to our knowledge, there has been no previous investigation targeting the association between tau and local brain age. Regarding cognition, global brain age has shown associations with cognitive measures^1–3^, although these are modest in strength and more pronounced in disease samples^1^,. Local brain age prediction may offer perspectives in this regard. Here, we examined the relationship between local brain aging patterns and performance measures derived from the Rey’s Auditory-Verbal Learning Test (RAVLT)^50^ in MCI, AD, and CN groups. RAVLT has been successfully used for early detection of MCI-related impairments in learning and memory and prediction of progression from MCI to AD that is comparable to MRI biomarkers^51,52^.

## 2. Materials and Methods

### 2.1. Datasets

T1-weighted brain MRI scans were collected from multiple public sources for developing and evaluating the brain age prediction model. These scans were obtained as part of studies that had been reviewed and approved by local ethics committees. From each dataset, we used data of adult participants (chronological age greater than or equal to 20 years). A brief description of each dataset, including the details of the initial data selection criteria employed in the present study, is provided in section S1.1. in Supplementary materials and methods. The used datasets include the Alzheimer’s Disease Neuroimaging Initiative (ADNI), Cambridge Centre for Aging and Neuroscience (Cam-CAN), Center for Biomedical Research Excellence (COBRE), Dallas Lifespan Brain Study (DLBS), Information eXtraction from Images (IXI), Nathan Kline Institute-Rockland Sample (NKI-RS), Neuroimaging in Frontotemporal Dementia (NIFD), Open Access Series of Imaging Studies (OASIS), Southwest University Adult Lifespan Dataset (SALD), and the UK Biobank (UKB) dataset. Demographic characteristics of the final set of participants from each dataset are summarized in Table S1 and depicted in Fig. S1 and S2. Note that the final sample described in the Supplementary Materials is derived by applying further data selection and partitioning steps detailed in Section 2.3 in the manuscript.

### 2.2. Data preprocessing

#### 2.2.1. MR image preprocessing and quality control

In-house preprocessing of the T1-weighted MRI scans included conversion from DICOM to NIfTI format, bias correction, spatial normalization to MNI152 space, resampling the images to 1 mm^3^ isotropic voxel resolution using 4th degree B-spline interpolation, and skull-stripping using the SPM12 toolbox (version 6685, https://www.fil.ion.ucl.ac.uk/spm/software/spm12/)^53,54^ and custom-made scripts running on MATLAB R2015a (MathWorks Inc., Natick, MA, United States). This was followed by manual review of the images to filter out scans where preprocessing failed. Spatial normalization to MNI standard space using SPM resulted in images with a size of 157 × 189 × 156 voxels. Voxel intensity values were standardized by subtracting the mean intensity value of the image and then dividing by the standard deviation. Subsequently, intensity values outside the [−1, 2.5] interval were clipped to the interval edges^39^.

Image quality was characterized by their Euler number^55^. The Euler numbers were obtained by running the FreeSurfer v7.1.1 software package^56^. Based on previous recommendations^57^, we transformed the Euler numbers using the following formula to obtain a more normalized distribution:

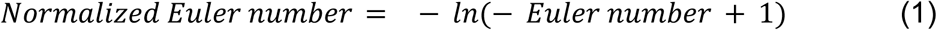

Normalization was performed for each dataset separately, across all groups in the given dataset. Then, images whose normalized Euler number was at or more than 1.5 inter quartile range (IQR) points below the first quartile of the data distribution were marked as outliers and removed from further analyses. The number of removed images ranged from 0 to 15 (0% to 2.63%) across datasets.

#### 2.2.2. Tau PET processing methods

We used tau PET data from ADNI-3 processed by UC Berkeley to investigate the relationship between local brain age and regional tau uptake in the temporal lobe. The tau PET data processing pipeline developed by the UC Berkeley team^58,59^ depends on a contemporaneous native space MRI that is segmented and parcellated with FreeSurfer v7.1.1. After using SPM12 to coregister each tau scan to the bias-corrected T1w MRI created by FreeSurfer, intensity values were normalized with reference to an inferior cerebellar gray matter region defined using the SUIT template^60^. The standardized uptake value ratio (SUVR) was then calculated within each FreeSurfer Desikan-Killiany region (left, right and volume-weighted bilateral) and a temporal meta-region of interest (ROI). We restricted our analyses to the temporal meta-ROI encompassing the bilateral entorhinal cortex, amygdala, parahippocampal, fusiform, inferior temporal and middle temporal cortical regions^58,59^, in line with previous results showing that this temporal meta-ROI shows the highest discriminative accuracy when separating AD from non-AD dementia^46^. The meta-ROI is the volume-weighted average of the individual regions and was not modelled as a single region during the analysis. We used data from the 6 mm resolution dataset containing SUVRs from FTP tracer. Tau data was corrected for partial volume effects using the Geometric Transfer Matrix (GTM) approach^61–64^. We only used data that passed quality control based on visual inspection of the FreeSurfer segmentation and MR/PET registration^59^.

### 2.3. Data partitioning

The preprocessed T1-weighted brain MRI scans from the Cam-CAN, DLBS, IXI, NKI-RS, SALD, and UKB datasets, as well as the images of CN subjects from the ADNI-2 dataset, were used for model training and validation. In the case of ADNI-2, subjects with a diagnosis other than CN on any occasion were omitted from the model development process. This ensured that the brain age prediction model was developed using scans from predominantly healthy participants. Euler numbers were normalized for each dataset separately, and outliers were identified based on the distribution of the normalized Euler numbers (see Section 2.2. for details), independently for each dataset, and were excluded from the model development and evaluation process. Then, the Cam-CAN, DLBS, IXI, NKI-RS, SALD, and UKB datasets were randomly divided into disjoint training and validation subsets with a 90:10 ratio, using stratified sampling based on chronological age and sex. The ADNI-2 images of CN subjects were divided into disjoint subsamples, resulting in training, validation, and test image sets with a 78:10:12 ratio, again using stratified sampling based on chronological age and sex. To avoid data leakage, all the images of ADNI-2 subjects with multiple MRI scans were assigned either to the training set, to the validation set, or to the test set. The resulting training set included 2938 images from 2425 subjects (1346 females, age range: 20 - 90 years, mean age ± standard deviation: 58.71 ± 17.60 years) and the validation set included 341 images from 275 subjects (155 females, age range: 20 - 88 years, mean age ± std: 59.10 ± 17.51 years; Table S1, Fig. S1). Only the images in the training and validation sets were used for model training and hyperparameter selection. As chronological age values varied in precision across datasets, all age values were rounded to whole years, and the age labels were divided by 100 during training. The ADNI-2 test set, including 100 images of 20 CN subjects (11 females, age range: 64 - 88 years, mean age ± std: 74.45 ± 6.44 years), constituted a small internal test sample to evaluate the generalization of the model in terms of age prediction performance to novel images of cognitively normal subjects. External test sets including the OASIS-3, NIFD, and COBRE datasets provided more stringent tests of generalization and the opportunity to investigate global and local brain aging in various disease states. Euler numbers were normalized for each dataset separately, considering all subject groups within the given dataset. As subjects in OASIS-3 and NIFD typically had multiple MRI scans, only the scan with the highest normalized Euler number (best image quality) was kept for each subject. Then, outliers were identified and removed based on the distribution of the normalized Euler numbers, independently for each dataset, again considering all subject groups within a particular dataset. Finally, subject groups within each dataset were matched in number of subjects, chronological age, and sex. For OASIS-3, three subject groups were defined based on the scores on the Clinical Dementia Rating Scale (CDR): CDR-0 (score equal to 0), CDR-0.5 (score equal to 0.5), and CDR-1 (score greater than or equal to 1), corresponding to CN, MCI, and AD, respectively^37^. For the NIFD and COBRE datasets, subject groups included individuals with bvFTD and SZ, respectively, in addition to CN control subjects. The distribution of age and sex in these subject groups are described in Table S1 and plotted in Fig. S2. No significant differences were found in chronological age between subject groups using an ANOVA in OASIS-3 (F = 0.1291, p = 0.88) or using independent two-tailed t-tests in NIFD (t = 0.27, p = 0.79) and COBRE (t = −1.23, p = 0.22).

To analyze longitudinal changes in local hippocampal brain age in stable and progressive MCI, we used data from the ADNI-2 and ADNI-3 datasets, ensuring that there was no overlap with the training/validation sets on the subject level. First, we selected progressive MCI subjects (pMCI) who converted from MCI to AD without later reversal to either MCI or normal and had a T1-weighted image at conversion and approximately 1 year and 2 years prior to conversion (3 images in total). At each visit when the images prior to the conversion were taken, the subjects were diagnosed with MCI. Second, we selected stable MCI subjects (sMCI) who had solely MCI diagnoses across all their visits and at least 4 T1-weighted images taken at consecutive 1-year intervals. They were matched in mean age across visits and sex to converters, and the first 3 of their images (taken at consecutive 1-year intervals) were used for the analysis. The resulting Hippocampal Brain Age dataset included 31 stable and 31 progressive MCI subjects (13 females/group), with 3 images from each subject. The distribution of age and sex in these subject groups are described in Table S1 and plotted in Fig. S2.

To investigate the relationship between local brain age and tau uptake in the temporal lobe, we used ADNI-3 data from subjects that were not included in the training and validation sets. For each subject, data from a single tau PET and T1-weighted MRI scan pair was kept, considering the scans that were the closest to each other in terms of acquisition date. If the difference between the scan acquisition dates exceeded 30 days, the subject was excluded from further analysis. A diagnostic label was acquired for each subject (CN/MCI/AD), considering the results of the diagnostic exam that was closest in time to the MRI acquisition date. If the difference between the diagnostic exam date and either the MRI or the PET acquisition date exceeded 180 days, the subject was excluded from further analysis. We also excluded subjects that had a diagnosis other than the current one on any other ADNI-3 diagnostic exams, and subjects that had MCI or dementia due to etiology other than Alzheimer’s disease. After outlier removal based on the normalized Euler number, an equal number of CN, MCI and AD subjects were selected for correlation analysis (N = 34 subjects, 12 females in each group). We refer to the resulting set of participants as the Tau PET dataset. Besides sex, chronological age was also matched across subject groups (Table S1, Fig. S2).

To analyze the relationship between local brain-PAD and RAVLT scores (Immediate and Percent Forgetting), we used data from the ADNI-2 and ADNI-3 datasets, ensuring that there was no overlap with the training/validation sets on the subject level. We used the available RAVLT scores from the visit corresponding to the MRI acquisition, removing subjects with missing RAVLT scores. Following Moradi et al.^51^, we excluded AD subjects with a Percent Forgetting score of 0, and any subject with negative Percent Forgetting. After further outlier removal based on the normalized Euler number, an equal number of CN (N = 188 subjects, 85 females), MCI (N = 188 subjects, 78 females) and AD subjects (N = 188 subjects, 78 females) were selected for correlation analysis, matched in sex and chronological age as closely as possible (Table S1, Fig. S2).

### 2.4. Model development

BrainAgeMap consists of two different deep neural network models; a base model for predicting local (regional) brain age and a head model for predicting global brain age, i.e. the overall estimated biological age of the brain (Fig. 1). These models correspond to 3D fully convolutional (3D-FCN) and convolutional (3D-CNN) networks, respectively. A brief description of the model development process is provided in the following two subsections.

**Fig. 1.**
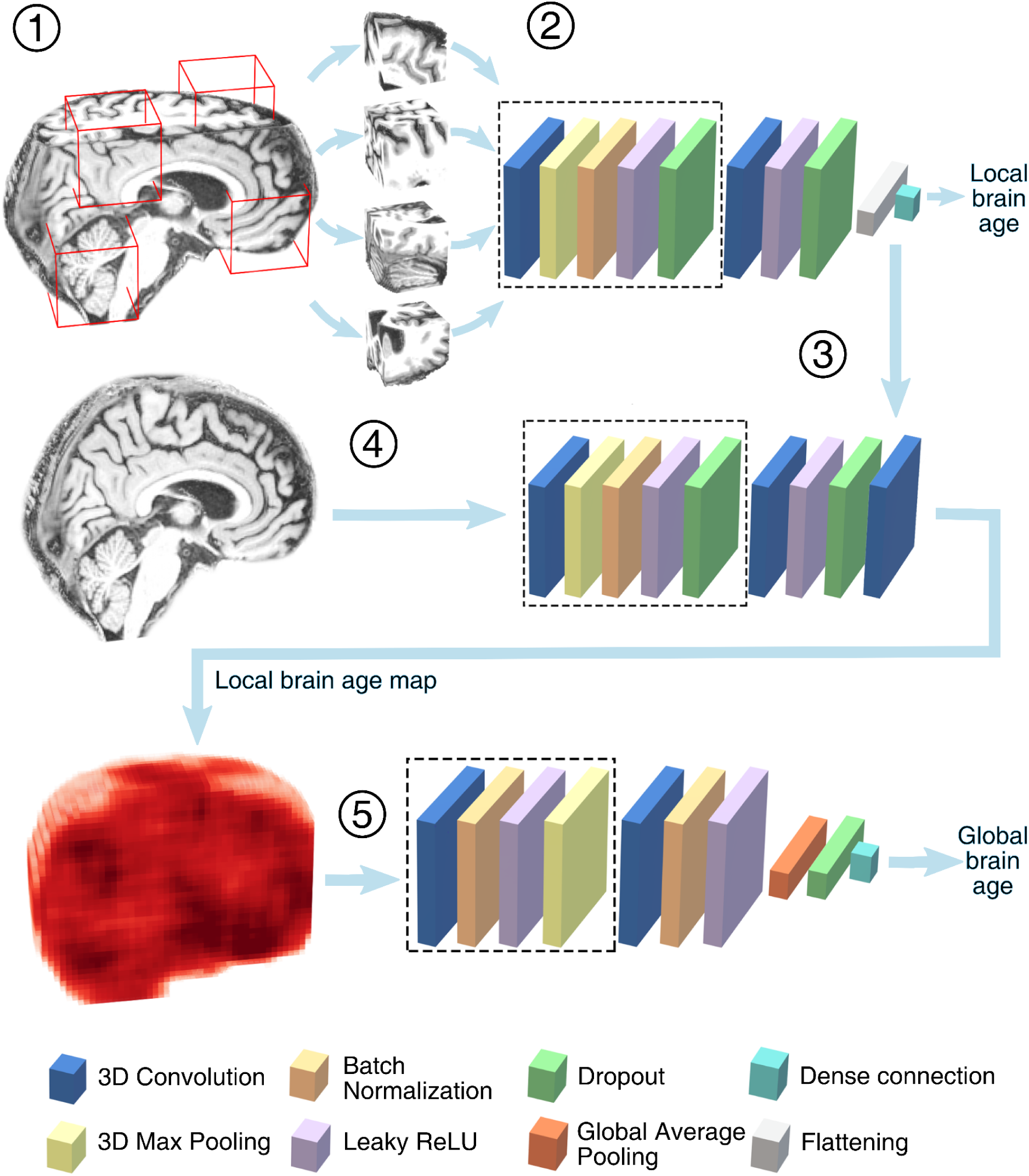
Schematic illustration of BrainAgeMap. Randomly selected cuboid patches are extracted from the preprocessed T1-weighted structural images (parts of the brain shown are only for illustration purposes) in the training set (1) and used for training a 3-D convolutional neural network (3D-CNN) base model to predict the chronological age of the participant (2). After being converted into a 3D fully convolutional network (3D-FCN) (3), the base model is applied to the whole image to obtain a local brain-predicted age map (4). The local brain-predicted age map is fed into a head model to obtain the global brain-predicted age of the participant: a 3D-CNN that uses the whole local brain-predicted age map as input (5). ReLU: rectified linear unit.

#### 2.4.1. Base model for local brain age prediction

The BrainAgeMap base model was trained to predict chronological age based on randomly selected cuboid patches^39^ of size 47 × 47 × 47 voxels, extracted from the preprocessed structural brain images in the training set. The base model is a 3D-CNN consisting of 5 convolutional blocks. The first 4 convolutional blocks consist of 3D convolution using VALID padding, max pooling, batch normalization^65^, and leaky rectified linear unit nonlinearity (leaky ReLU)^66^, and the last convolutional block consists of 3D convolution and leaky ReLU. The output of the last convolutional block was flattened and fed into a fully connected output layer consisting of a single unit with linear activation. For a full specification of the network, see Table S2. During training, dropout regularization^67^ (with dropout rates ranging between 0.1 and 0.5) was applied after each leaky ReLU nonlinearity. The weights of the hidden layers were initialized using He normal initialization^68^, and the weights of the output layer were initialized using Glorot uniform initialization^69^. The bias term in the fully connected output layer was initialized to 0. The model has 1,522,511 parameters, out of which 1,521,911 are trainable parameters.

The base model was trained for 100 epochs using the mean squared error (MSE) loss function and the Adam optimizer^70^ algorithm with a learning rate of 0.0001 and a batch size of 10. In each epoch, 300 random cuboid patches were extracted from each training set image and fed into the network in a random order. Data augmentation was applied by randomly changing brightness and contrast, and by adding Gaussian noise to the patches^39^. After training was finished, the model was converted into a 3D-FCN by discarding the flattening layer and converting the fully connected (dense) output layer into a 3D convolutional layer using the weights and bias term learned by the original dense layer^71^ (Table S2). This allowed the application of the network to 3D images of arbitrary size. For each training epoch, the model parameters were set to the values learned at the end of the epoch, and the 3D-FCN was applied to the validation set. During inference, each image was symmetrically padded, with the value of the voxel in the farthest corner of the image^39^, to a size of 203 × 205 × 202 voxels. Application of the 3D-FCN to the (padded) whole images resulted in heatmaps of size 40 × 48 × 39 voxels corresponding to local brain-predicted ages for each scan in the validation set. The mean absolute error (MAE) between the chronological age and the average local brain-predicted age was calculated for each epoch, and the parameter values corresponding to the epoch with the lowest validation set MAE were retained in the final model that was subsequently evaluated on the test sets (see Section 2.5.).

#### 2.4.2. Head model for global brain age prediction

The estimation of global brain age was based on a 3D-CNN model applied to the whole local brain age map. The network consists of a stack of 4 convolutional blocks. Each convolutional block consists of 3D convolution with 3 × 3 × 3 filters and SAME padding, batch normalization, and leaky ReLU non-linearity. The number of filters in the successive convolutional layers were [16, 32, 32, 64]. Each convolutional block is followed by max pooling with a pool size of 2 and a stride of 2, except for the last one which is followed by global average pooling. Dropout regularization (with a dropout rate of 0.5) was applied to the global average pooling layer during training. The output of the global average pooling layer is fed into a fully connected output layer consisting of a single unit with linear activation (Table S3). He normal initialization was used in the hidden layers to initialize kernel weights, and Glorot uniform initialization was used to initialize the weights of the fully connected output layer. The bias term in the fully connected output layer was initialized to 0. The model has 97,841 parameters, out of which 97,553 are trainable.

The model was trained using the MSE loss and the Adam optimizer with a constant learning rate of 0.01 and a batch size of 10. Early stopping was applied with a patience of 20 epochs, i.e. the model was evaluated on the validation set after each epoch and training was stopped when the validation loss had not decreased for the last 20 epochs. The learned parameter values corresponding to the epoch with the lowest validation loss were retrieved, and the model was used for evaluation on the test sets. The neural networks were implemented in Python 3.8.10 using TensorFlow^72^ 2.4.1.

### 2.5. Model evaluation

The following sections describe the methods used to evaluate the local and global brain age predictions of the trained BrainAgeMap model. Statistical analyses regarding the comparison of local and global brain age between subject groups (section 2.5.1.) and the relationship between local brain age and tau uptake (section 2.5.3.) and RAVLT summary scores (section 2.5.4.) were performed using the Pingouin^73^ 0.5.3. statistical package for Python 3. Linear mixed effects modelling and its visualization for the longitudinal analysis (section 2.5.2.) were performed in RStudio^74^ (version 2025.05.0+496) using R^75^ (version 4.5.0) and the tidyverse^76^, ggplot2^77^, and dplyr^78^ packages.

#### 2.5.1. Comparing local and global brain age between subject groups

The final FCN base model was evaluated on the internal ADNI-2 test set and then used to compare local brain-PAD between healthy control subjects and subjects with mild cognitive impairment and Alzheimer’s disease (OASIS-3 dataset), frontotemporal dementia (NIFD dataset), and schizophrenia (COBRE dataset). The base model was applied to each image in the test sets to obtain local (regional) brain age maps. We addressed a particular bias that is commonly observed in brain age prediction, namely the over- and underestimation of the age of younger and elderly subjects, respectively^79^, by applying a correction procedure to brain-predicted ages. The correction method included modelling the relationship between brain-predicted and chronological age using the validation set:

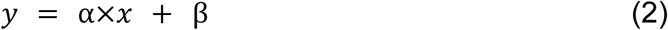

where x is the chronological age, *y* is the original brain-predicted age, and α and β are the estimated coefficient and intercept term, respectively. Then, brain-predicted age on the test sets was corrected using the formula:

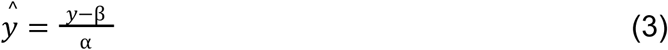

where *y^^^* is the corrected brain-predicted age, as in Peng et al.^13^ This correction procedure was applied to each voxel separately. These corrections were performed for all test set evaluations, including the longitudinal analysis of local brain age (section 2.5.2.), and the analysis of the relationships between local brain age and tau uptake (section 2.5.3.) and RAVLT summary scores (section 2.5.4.).

After correction, local brain-PAD maps were obtained for each scan by subtracting the chronological age of the participant from each brain-predicted age value in the brain age map. These local brain-PAD maps were compared between each patient group and the corresponding healthy control group in each test set using voxel-wise analyses of covariances (ANCOVAs) with the normalized Euler number and chronological age as covariates. Statistical comparisons were restricted to voxels within the intracranial mask of the SPM toolbox (tpm/mask_ICV.nii). Correction for multiple comparisons was performed by applying the Benjamini-Yekutieli procedure that controls the false discovery rate (FDR) at level α under arbitrary dependence assumptions^80^. In our analyses, the value of α was set to 0.05. To visualize the results, grand average local brain-PAD maps were obtained by averaging the brain-PAD maps across subjects in each subject group separately. The values of voxels where no significant difference was observed between the subject groups were set to 0. Then, the grand average brain-PAD map of the subject group was subtracted from the grand average brain-PAD map of the patient group. Finally, the resulting difference brain-PAD map was upsampled using the ‘zoom’ function of the multi-dimensional image processing package (scipy.ndimage) of the SciPy ecosystem (https://scipy.org/) with zero-order spline interpolation and overlaid on a structural template image derived by averaging the T1-weighted images in the patient group using the spm_imcalc function.

The head model was also applied to each image in the test sets to obtain global brain-predicted ages. These brain-predicted ages were also corrected based on the global brain-predicted ages on the validation set, using the procedure described above. Model accuracy was estimated using the correlation coefficients between the predicted brain age and the chronological age (Spearman’s ρ; partial correlation using the normalized Euler number as a covariate, with 95% parametric confidence intervals (CI) around the correlation coefficient estimated using the Fisher transformation), MAE, root mean squared error (RMSE), the coefficient of determination (R2), and the mean and standard deviation of the brain-PAD, for each subject group separately. Group differences in global brain-PAD (global brain age predicted by the head model minus chronological age) between patient and control groups were assessed by ANCOVAs with the normalized Euler number and chronological age as covariates. Partial eta-squared (η_p_ ^2^) effect sizes are reported. Post hoc tests (OASIS-3) were performed using pairwise ANCOVAs and *p* values were adjusted using Bonferroni correction.

#### 2.5.2. Longitudinal analysis of hippocampal brain age

The FCN base model was used to obtain bias corrected brain age maps for each image of the sMCI and pMCI subjects in the Hippocampal Brain Age dataset. A hippocampal mask was determined with reference to the Neuromorphometrics atlas of SPM12. Maximum probability tissue labels were derived from the “MICCAI 2012 Grand Challenge and Workshop on Multi-Atlas Labeling” (https://masi.vuse.vanderbilt.edu/workshop2012/index.php/Challenge_Details) and were provided by Neuromorphometrics, Inc. (http://Neuromorphometrics.com/) under academic subscription. The Neuromorphometrics label map was downsampled using zero-order spline interpolation to match the size of the local brain age maps, and then local brain age values were averaged across voxels corresponding to the left and right hippocampus, for each image separately. The mean hippocampal brain-PAD values were obtained by subtracting the chronological age from the mean brain-predicted age. These values were entered into a linear mixed model with group (stable versus progressive), visit number (1–3) and their interaction as fixed effects, and random intercepts and slopes for each participant. The model additionally included chronological age, and scan quality as assessed by Euler number as covariates.

#### 2.5.3. Relationship between local brain age and tau uptake

The FCN base model was used to obtain bias corrected brain-predicted age maps for each image in the Tau PET dataset. These brain age maps were then transformed to native space using a custom MATLAB script and SPM12. FreeSurfer v7.1.1. was used to segment the T1-weighted scans based on the Desikan-Killiany atlas, and the segmentation masks were converted to native space using the FreeSurfer “mri_label2vol” command. Brain-predicted age values were averaged within the temporal meta-ROI encompassing the bilateral entorhinal cortex, amygdala, parahippocampal, fusiform, inferior temporal and middle temporal cortical regions. After subtracting chronological age, individual temporal meta-ROI brain-PAD values were correlated with the temporal meta-ROI tau PET SUVRs (corrected for partial volume effects) using partial Spearman correlations with the normalized Euler number and chronological age as covariates, separately in the CN, MCI, and AD subject groups. *P* values were corrected for multiple comparisons using the Bonferroni method.

#### 2.5.4. Relationship between local brain age and RAVLT summary scores

In the present study, two different summary scores, Immediate and Percent Forgetting, were calculated from the raw RAVLT scores, as in Moradi et al.^51^ RAVLT Immediate—a proxy for verbal learning—was calculated as the sum of scores from the first five consecutive trials (Trials 1-5). RAVLT Percent Forgetting—a proxy for delayed memory—was calculated as the score of Trial 5 minus the score of the delayed recall, divided by the score of Trial 5. The distribution of the RAVLT Immediate and Percent Forgetting summary scores are shown in Fig. S3 and S4, respectively. To investigate the relationship between local brain-PAD and the RAVLT summary scores, voxel-wise partial Spearman correlations were calculated in each subject group (CN/MCI/AD), with chronological age and the normalized Euler number as covariates. The Benjamini-Yekutieli procedure^80^ was used to correct for multiple comparisons. The resulting maps, showing the topographical distribution of Spearman’s correlation coefficient, were upsampled and overlaid on the structural template of the corresponding subject group. The value of the coefficient was set to zero in voxels where the correlation was not significant after correction for multiple comparisons. For a short description of RAVLT, see section S1.3. in Supplementary materials and methods.

## 3. Results

The FCN base model achieved the lowest validation MAE of 7.47 years (across all voxels) after 66 epochs. The CNN head model used for global brain age prediction obtained a validation MAE of 3.62 years. In all subsequent evaluations, we used the FCN base model for local brain age prediction and the CNN head model for global brain age prediction. We refer to this model pair as BrainAgeMap. BrainAgeMap generalized well in terms of global brain-predicted age to the CN ADNI-2 test set (Spearman’s ρ = 0.92, *p* < 0.001, R2 = 0.73, MAE = 2.61 years, RMSE = 3.29 years; Fig. S5), and revealed meaningful patterns of local and global brain aging in various disease states, as detailed in the following subsections.

### 3.1. Local and global brain age in neurodegenerative and neuropsychiatric disease states

The application of the FCN base model to the OASIS-3 test set revealed significantly greater local brain-PAD in temporal lobe regions in MCI compared to CN, with a somewhat more pronounced effect in the left than in the right hemisphere (Fig. 2, Fig. S6). Significant differences were observed bilaterally in the hippocampus, entorhinal cortex, parahippocampal gyrus, amygdala, and in the left fusiform gyrus. In addition, significant differences were also found in the lateral ventricle, thalamus, caudate, and putamen regions. The comparison of AD subjects to controls (Fig. 2, Fig. S6) revealed significantly higher local brain-PAD in a more extensive region in the temporal lobe bilaterally that also extended to posterior regions encompassing the inferior and middle occipital gyri. Significant differences were also found in the posterior and middle cingulate regions, in the lateral and third ventricles and in subcortical structures including the thalamus and caudate. Comparison of MCI and AD subjects revealed no significant differences in local brain-PAD.

**Fig. 2.**
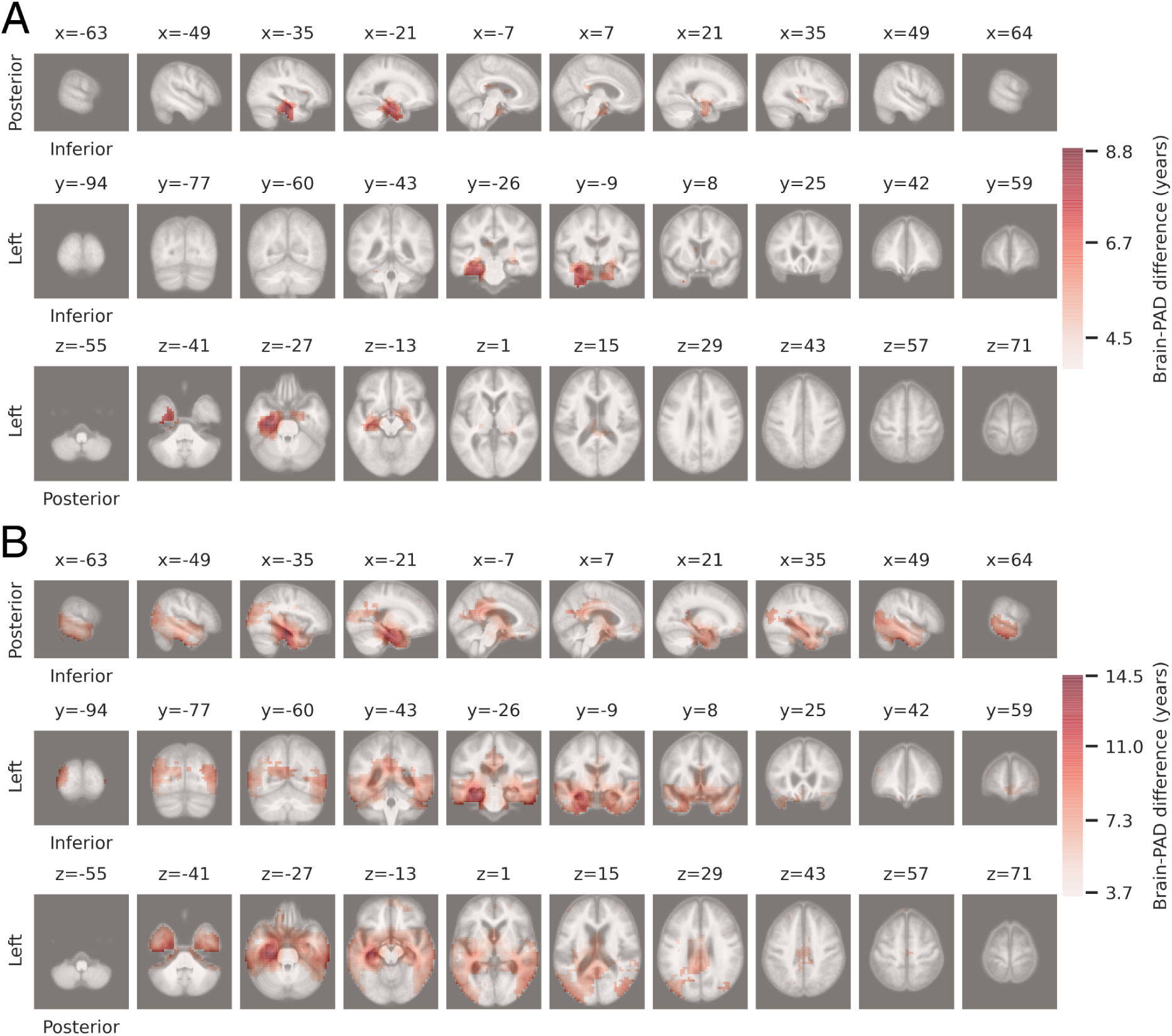
Grand average brain-PAD difference maps in the OASIS-3 test set. A) Grand average brain-PAD difference maps showing increased local brain-PAD in subjects with mild cognitive impairment (MCI) compared to cognitively normal control (CN) subjects. The difference in local brain-PAD (brain-predicted minus chronological age) between the MCI and CN groups is shown for voxels where the difference was significant after correction for multiple comparisons. B) Grand average brain-PAD difference maps showing increased local brain-PAD in subjects with Alzheimer’s disease (AD) compared to CN subjects.

Global brain-predicted age and chronological age in the OASIS-3 dataset were strongly correlated in the CN group (Spearman’s ρ = 0.72, CI [0.56,0.83], R2 = 0.53, MAE = 4.19 years, RMSE = 5.22 years), and moderately strong correlations were observed in the MCI (Spearman’s ρ = 0.60, CI [0.4,0.75], R2 = 0.34, MAE = 4.36 years, RMSE = 5.63 years) and AD groups (Spearman’s ρ = 0.47, CI [0.23,0.66], R2 = 0.33, MAE = 5.93 years, RMSE = 7.17 years; Fig. 3). All correlations were significant at *p* < 0.001. Global brain-PAD differed significantly between subject groups (F = 8.03, DF = 2, η_p_ ^2^ = 0.09, *p* < 0.001). Post hoc tests revealed significantly higher brain-PAD in the MCI (mean brain-PAD ± standard deviation =1.56 ± 5.46 years; F = 7.51, DF = 1, η_p_ ^2^ = 0.07, *p* = 0.022) and AD (mean brain-PAD ± standard deviation = 2.60 ± 6.74 years; F = 12.52, DF = 1, η_p_ ^2^ = 0.11, *p* = 0.002) groups than in the CN group (mean brain-PAD ± standard deviation = −0.87 ± 5.19 years). The difference between the MCI and AD groups was not significant (F = 2.65, DF = 1, η_p_ ^2^ = 0.02, *p* = 0.32).

**Fig. 3.**
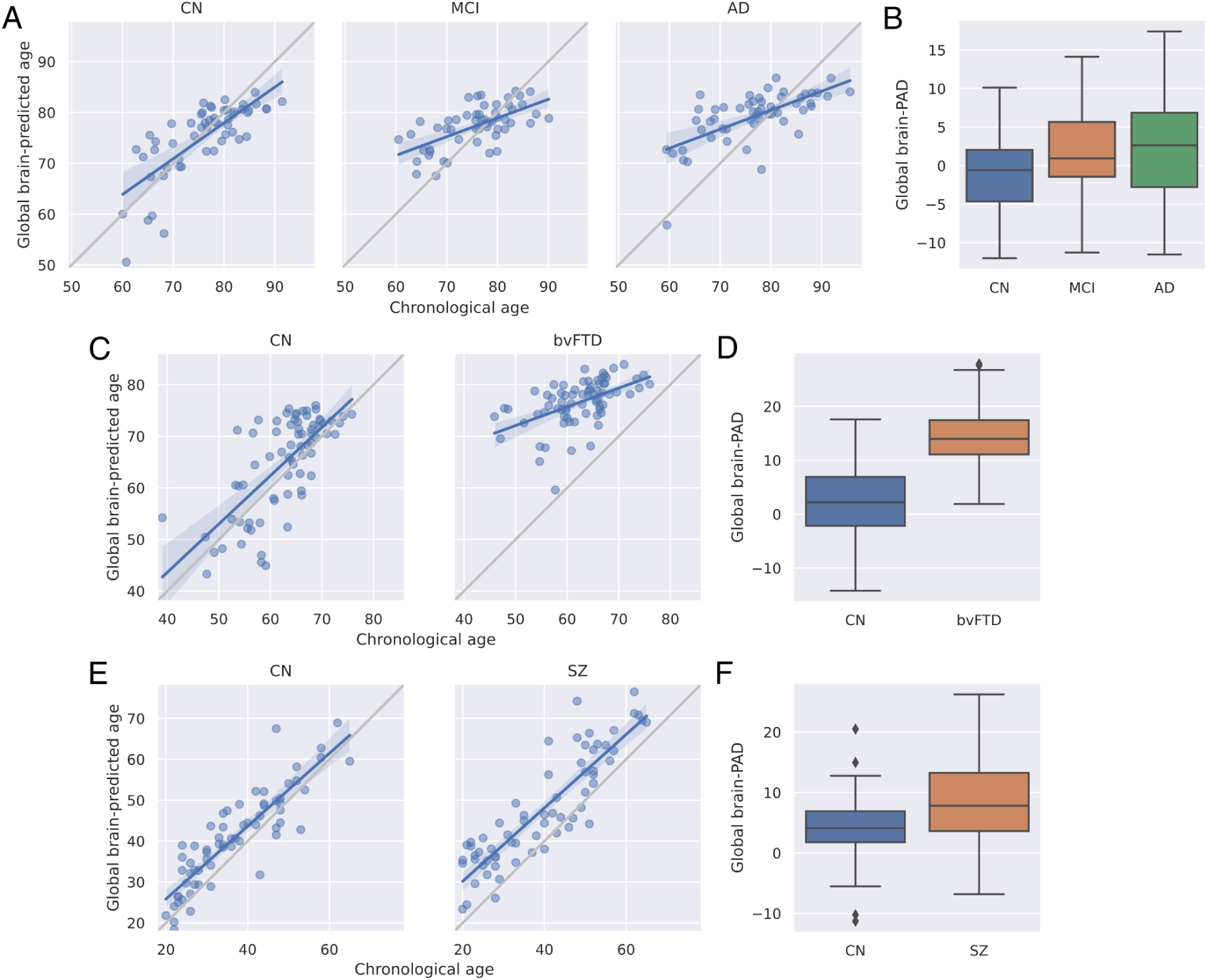
Global brain-predicted age in the test sets. A) Correlation between chronological age and global brain-predicted age (years) in cognitively normal (CN) subjects and subjects with mild cognitive impairment (MCI) and Alzheimer’s disease (AD) in the OASIS-3 test set. Blue and gray lines correspond to the regression lines and the lines of identity, respectively. The shaded areas represent the 95% confidence intervals. B) Distribution of global brain-PAD (brain-predicted minus chronological age) in the OASIS-3 test set. Whiskers represent 1.5 interquartile ranges, horizontal lines inside the boxes denote the median. C) Correlation between chronological age and global brain-predicted age in CN subjects and subjects with behavioral-variant frontotemporal dementia (bvFTD) in the NIFD test set. D) Distribution of global brain-PAD in the NIFD test set. E) Correlation between chronological age and global brain-predicted age in CN subjects and subjects with schizophrenia (SZ) in the COBRE test set. F) Distribution of global brain-PAD in the COBRE test set.

With regard to frontotemporal dementia (the NIFD dataset), significantly greater local brain-PAD was found in bvFTD subjects compared to healthy controls in virtually every brain region. However, the most pronounced increase in local brain-PAD was observed in anterior brain regions in the frontal and temporal lobes (Fig. 4, Fig. S7).

**Fig. 4.**
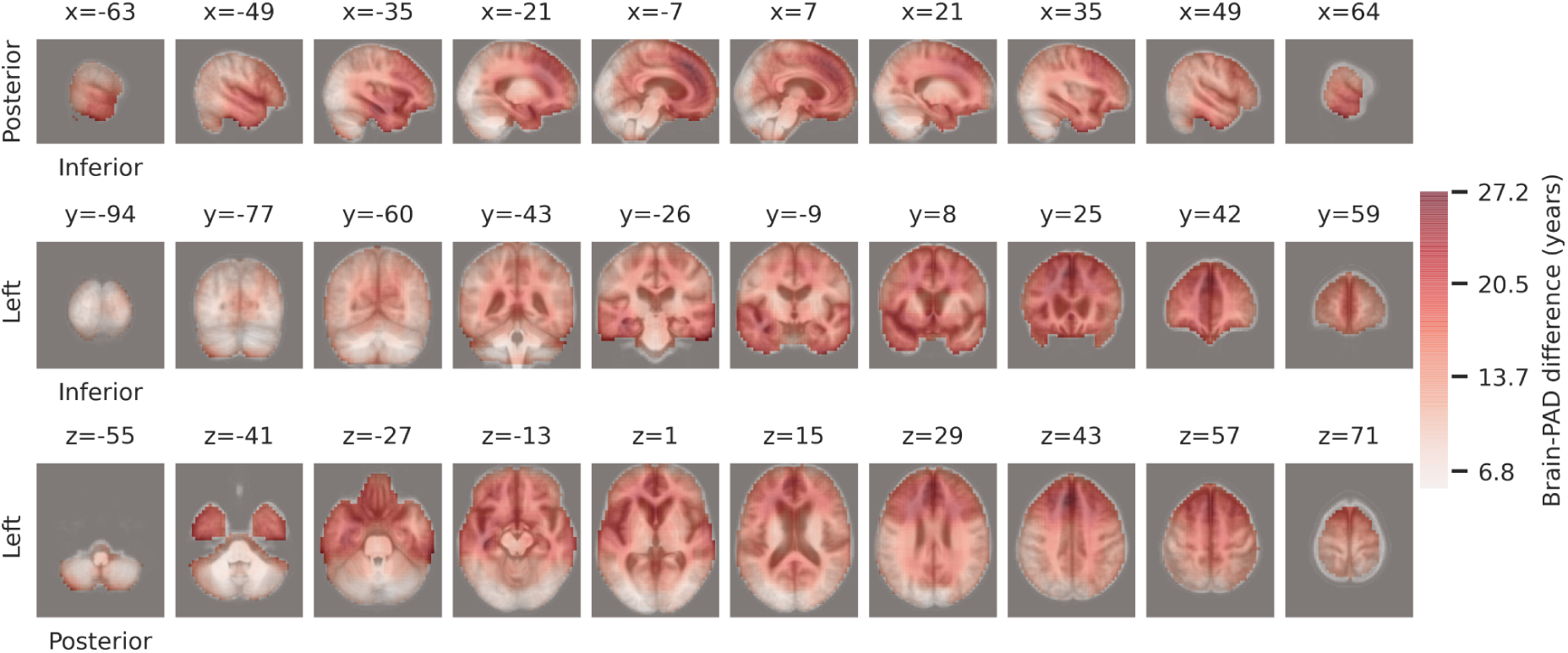
Grand average brain-PAD difference maps showing increased local brain-PAD in subjects with behavioral-variant frontotemporal dementia (bvFTD) compared to cognitively normal control subjects (CN) in the NIFD dataset. The difference in local brain-PAD (brain-predicted age minus chronological age) between the bvFTD and CN groups is shown for voxels where the difference was significant after correction for multiple comparisons.

Global brain-predicted age and chronological age were strongly correlated in the CN group (Spearman’s ρ = 0.66, CI [0.5,0.77], R2 = 0.03, MAE = 5.47 years, RMSE = 6.90 years), and a somewhat weaker correlation was observed in the bvFTD group (Spearman’s ρ = 0.50, CI [0.31,0.66], R2 = −4.84, MAE = 14.21 years, RMSE = 15.24 years; Fig. 3). All correlations were significant at *p* < 0.01. Global brain-PAD was significantly higher in the bvFTD group (mean brain-PAD ± standard deviation = 14.21 ± 5.54 years) than in the CN group (mean brain-PAD ± standard deviation = 2.19 ± 6.59 years; F = 94.87, DF = 1, η_p_ ^2^ = 0.40, *p* < 0.001).

The investigation of local brain age in the COBRE dataset revealed significantly higher local brain-PAD in SZ subjects than in healthy controls in an extensive region covering the prefrontal cortex and precentral gyrus bilaterally, and in bilateral temporal, insular, and opercular cortical areas (Fig. 5, Fig. S8). Significant differences were also observed in more posterior areas, namely in the posterior cingulate and precuneus bilaterally, and in the right angular and middle occipital gyrus.

**Fig. 5.**
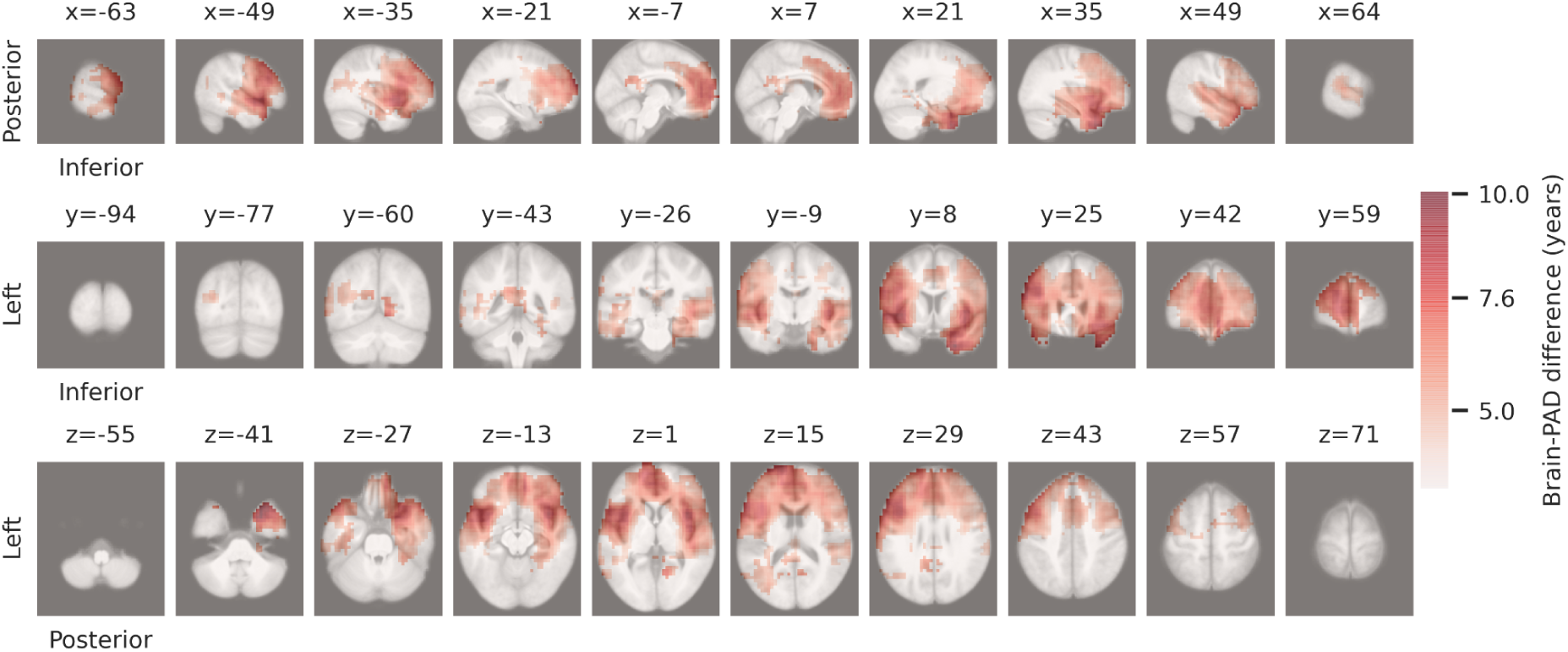
Grand average brain-PAD difference maps showing increased local brain-PAD in subjects with schizophrenia (SZ) compared to cognitively normal control subjects (CN) in the COBRE dataset. The difference in local brain-PAD (brain-predicted age minus chronological age) between the SZ and CN groups is shown for voxels where the difference was significant after correction for multiple comparisons.

Regarding global brain age in the COBRE dataset, strong correlations were observed between global brain-predicted age and chronological age in the CN (Spearman’s ρ = 0.90, CI [0.83,0.94], R2 = 0.63, MAE = 5.61 years, RMSE = 6.86 years) and in the SZ (Spearman’s ρ = 0.88, CI [0.81,0.92], R2 = 0.37, MAE = 8.70 years, RMSE = 10.44 years; Fig. 3) groups. All correlations were significant at *p* < 0.001. Global brain-PAD was significantly higher in the SZ group (mean brain-PAD ± standard deviation = 8.19 ± 6.53 years) than in the CN group (mean brain-PAD ± standard deviation = 4.00 ± 5.62 years; F = 17.73, DF = 1, η_p_ ^2^ = 0.12, *p* < 0.001).

### 3.2. Hippocampal brain age in stable and progressive mild cognitive impairment

A linear mixed model predicting local brain-PAD (see Table S4) revealed that compared to stable MCI participants, progressive MCI participants had higher average hippocampal brain-PAD (group main effect: *β* = 7.21, *p* < 0.001; Fig. 6.), and showed a steeper increase in brain-PAD across visits (visit number X group interaction: *β* = 1.00, *p* = 0.001, Fig. 6B). Importantly, post-hoc comparisons revealed that brain-PAD was consistently significantly higher for progressive than stable MCI participants (*p* < 0.001, Bonferroni-corrected, across all visits; Visit1: *b* = −8.16, 95% CI [−10.7,-5.63], *t*(5.63) = −6.453; Visit2: *b* = −9.34, 95% CI [−11.8,-6.91], *t*(63.1) = −7.675; Visit3: *b* = −10.16, 95% CI [−12.6,-7.74], *t*(59.9) = −8.407). We also observed a significant negative main effect of chronological age (*β* = – 0.32, *p* < 0.001), but not scan quality (*β* = – 0.1, *p* = 0.80) on brain-PAD in this model.

**Fig. 6.**
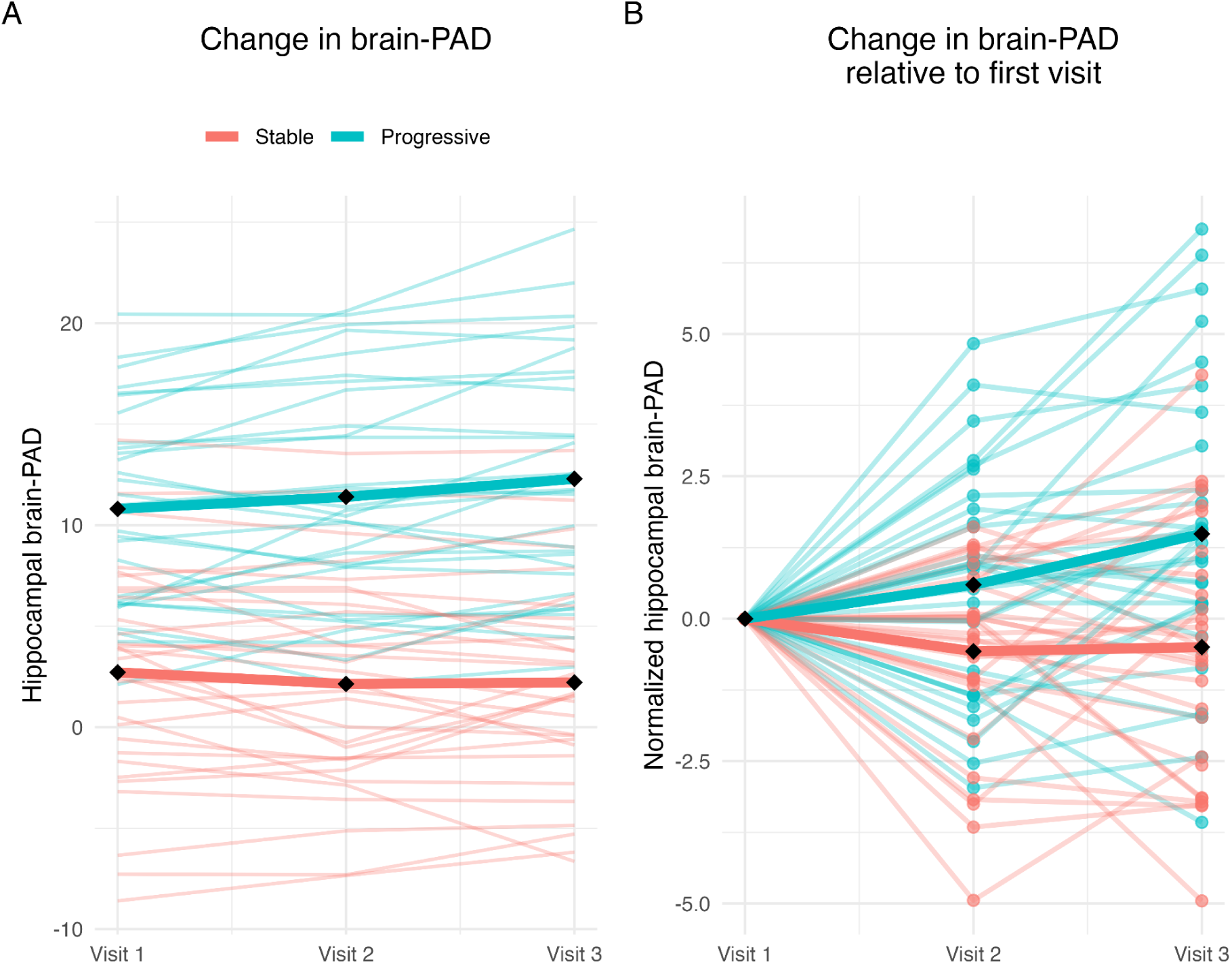
Individual (thin lines) and mean trajectories (thick lines) of longitudinal change in hippocampal brain-PAD (brain-predicted minus chronological age) across three visits for participants with stable (red) and progressive (blue) mild cognitive impairment. Raw hippocampal brain-PAD values (A) are normalized to baseline (Visit 1) in (B) to visualize individual differences in change.

A linear mixed model predicting global brain-PAD (see Table S5) revealed a similar pattern of results. Comparing standardized estimates between models revealed that the main effect of group in the local brain-PAD model (*β = 1.32, 95% CI [0.99,1.66])* was significantly stronger than in the global brain-PAD model (*β = 0.79, 95% CI [0.48,1.11])* Δβ = 0.53, 95% CI [0.07,0.99], *z* = 2.25, *p* = 0.024. In addition, the main effect of age in the local brain-PAD model (*β* = −0.35, 95% CI [−0.53,-0.18]) was significantly weaker than in the global brain-PAD model (*β* = −0.77, 95% CI [−0.92,-0.61]), Δβ = 0.41, 95% CI [0.18,0.65], *z* = 3.46, *p* = 0.001. No other effects differed between the two models.

### 3.3. Local brain age and tau uptake in the temporal lobe

Tau uptake (SUVR) showed a significant positive correlation with brain-PAD in the temporal meta-region of interest in AD subjects (mean brain-PAD ± standard deviation = 8.64 ± 7.74 years, mean SUVR ± standard deviation = 1.94 ± 0.65, Spearman’s ρ = 0.45, CI [0.12,0.69], *p* = 0.031; Fig. 7). A tendency toward a similar relationship was observed in the case of MCI subjects (mean brain-PAD ± standard deviation = 1.86 ± 5.05 years, mean SUVR ± standard deviation = 1.41 ± 0.38, Spearman’s ρ = 0.41, CI [0.08,0.67], *p* = 0.056). The association between SUVR and brain-PAD was not significant in the CN group (mean brain-PAD ± standard deviation = 0.93 ± 6.41 years, mean SUVR ± standard deviation = 1.21 ± 0.11, Spearman’s ρ = 0.004, CI [0.35,0.35], *p* = 1.00).

**Fig. 7.**
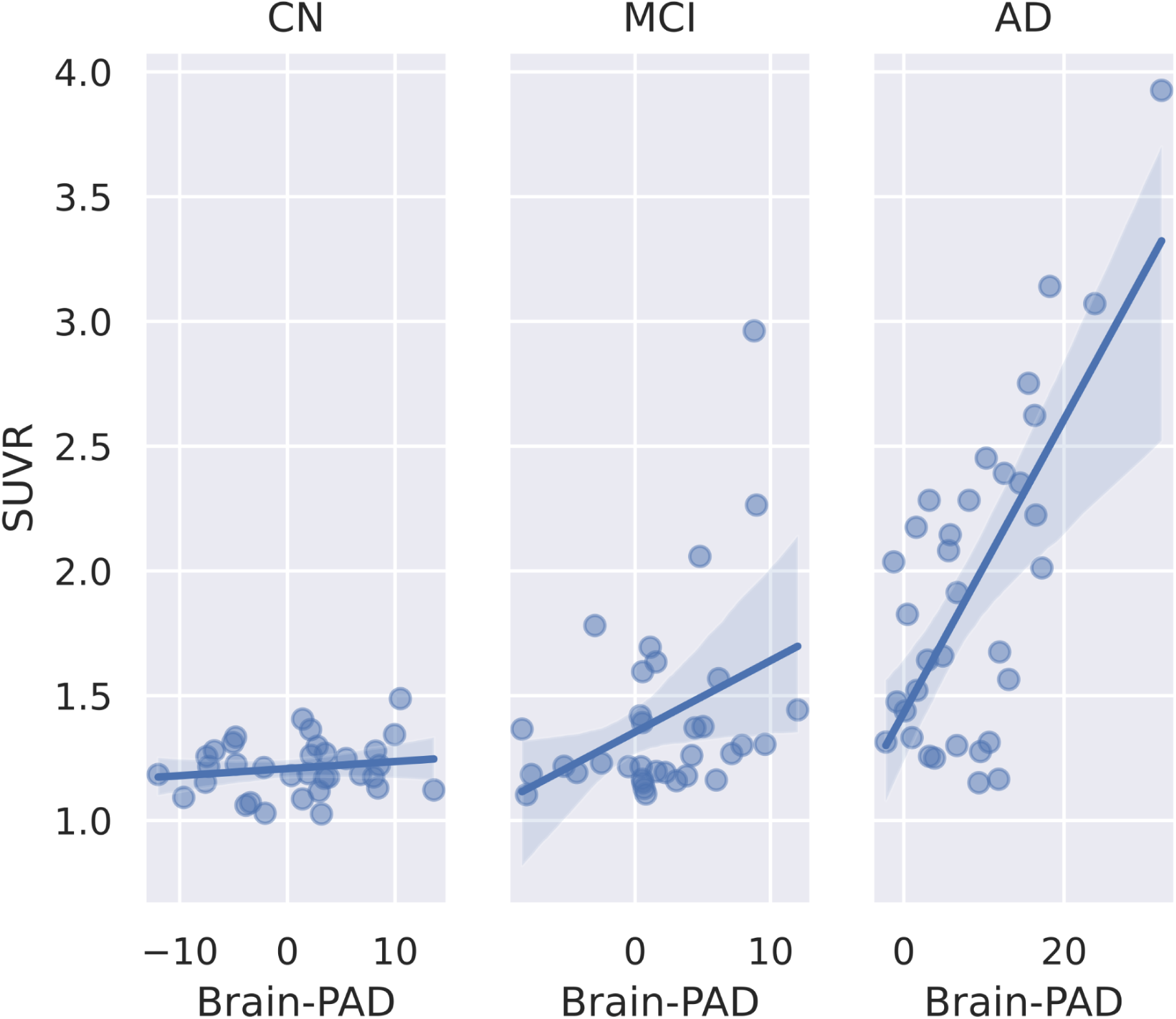
Relationship between brain-PAD (brain-predicted minus chronological age) and tau PET standardized uptake value ratio (SUVR) after partial volume correction in the temporal lobe in cognitively normal (CN) subjects, and subjects with mild cognitive impairment (MCI) and Alzheimer’s disease (AD). Blue lines correspond to the regression lines, and the shaded areas represent the 95% confidence intervals.

### 3.4. Local brain age and RAVLT summary scores

A significant negative correlation between local brain-PAD and RAVLT Immediate was observed in the MCI subjects in a bilateral temporal lobe region encompassing the hippocampus, entorhinal area, parahippocampal and fusiform gyrus. This area was more extensive in the left than in the right hemisphere. Additionally, significant correlations were observed in smaller patches in several cortical and subcortical areas (Fig. 8). Significant negative correlations were also observed in AD subjects in large cerebral areas, again with a greater extent in the left hemisphere (Fig. 8). No significant relationship was observed in the CN subjects.

**Fig. 8.**
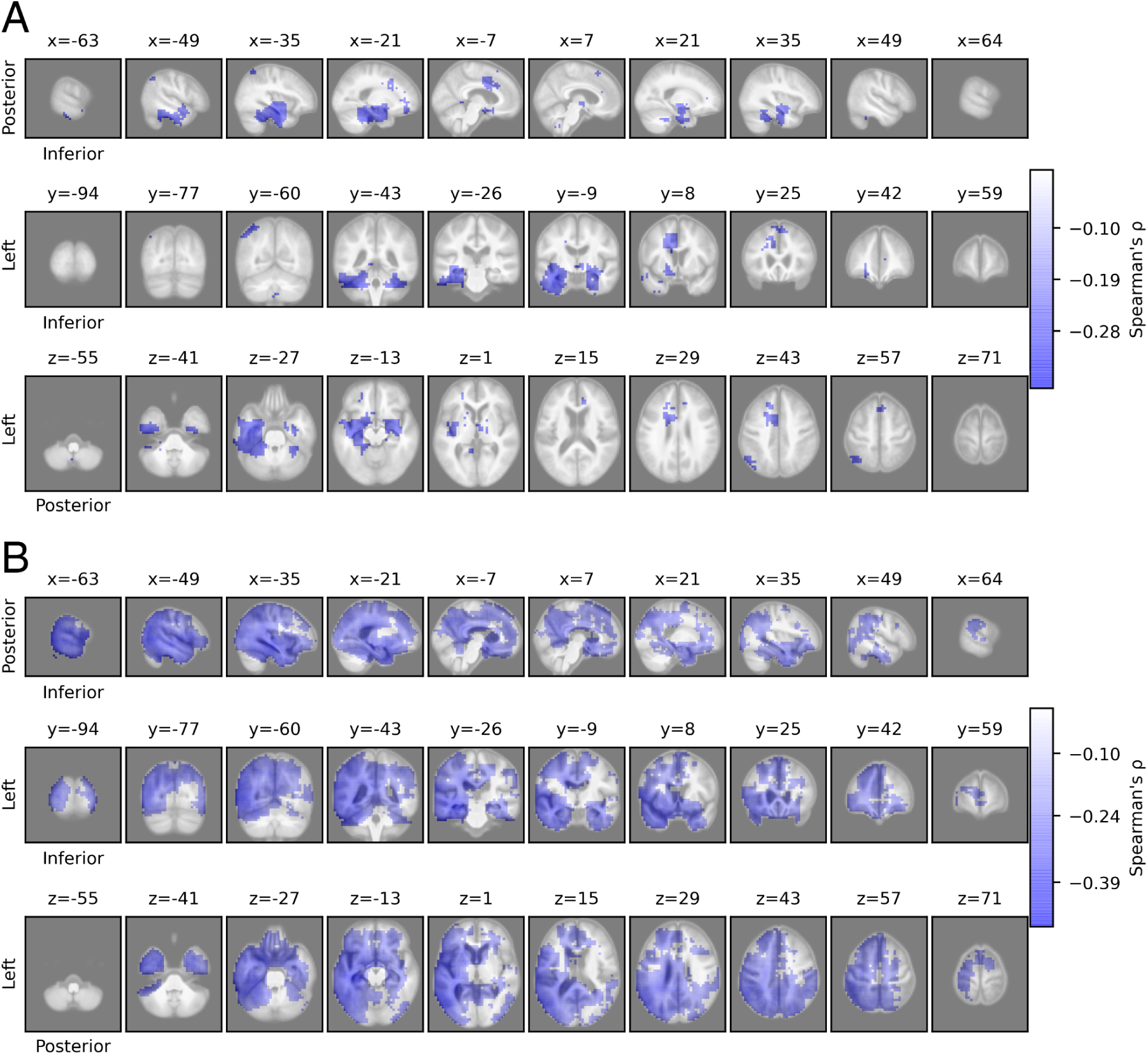
Associations between local brain-PAD and episodic memory function. Topographical distribution of the correlation coefficient (Spearman’s ρ) when assessing the correlation between local brain-PAD (brain-predicted minus chronological age) and the Immediate score in Rey’s Auditory Verbal Learning Test (RAVLT) in subjects with mild cognitive impairment (A) and Alzheimer’s disease (B). The coefficient is shown only for voxels where the correlation was significant after correction for multiple comparisons.

Regarding RAVLT Percent Forgetting, we observed a significant positive relationship with local brain-PAD mainly in the left temporal lobe regions of AD subjects (Fig. S9). However, this result should be treated with caution due to the highly skewed distribution of the summary score (Fig. S4). No significant correlations were observed in the CN and MCI groups.

## 4. Discussion

In the present study, we introduced a novel deep learning model, called BrainAgeMap, to estimate global and local brain age simultaneously based on T1-weighted MRI scans. Using this method, we investigated the brain aging patterns associated with various brain diseases. In line with previous observations of advanced brain aging in mild cognitive impairment and Alzheimer’s disease^10,81–83^, we found significantly higher global brain-PAD in MCI and in AD compared to healthy controls. This advanced global brain age was accompanied by a significantly larger local brain-PAD that was most pronounced in medial temporal lobe structures in MCI and spread over larger areas encompassing more posterior cortical as well as subcortical structures in AD. This overall pattern of advanced local brain aging is consistent with the anatomical abnormalities identified by volumetric MRI studies, namely gray matter atrophy beginning in the medial temporal lobe and progressing to an extensive set of cortical and subcortical regions in AD^84–86^. Global brain-PAD was remarkably higher in behavioral variant frontotemporal dementia than in healthy controls, which supports previous findings of advanced brain aging in FTD that is even more pronounced than in AD^36,49^. At the local level, we observed significantly increased brain-PAD practically throughout the entire brain, although the greatest increase occurred in frontal and temporal lobe areas. Neuroimaging studies indicate that disproportionate frontal, insular, or anterior temporal lobe atrophy may be helpful in distinguishing bvFTD from healthy aging and other dementias^87^, although neurodegeneration in subcortical structures may precede this canonical cortical atrophy pattern^88^. Finally, our results revealed significantly higher global brain-PAD in SZ patients when compared to healthy controls, in line with several previous studies examining structural brain age prediction in schizophrenia^6,89–92^. We also found a significant increase in local brain-PAD, mainly in prefrontal, temporal, insular, and opercular cortical areas. There is considerable overlap between this set of regions and those identified in voxel-based morphometry studies investigating brain structural alterations in SZ^93–95^. A meta-analysis of cross-sectional studies including 4789 SZ patients showed that schizophrenia is characterized by decreased gray matter volume in frontotemporal clusters, the parietal cortex, and subcortical structures^95^. The available evidence also points to a progressive component in the brain structural abnormalities associated with schizophrenia, as a meta-analysis of longitudinal volumetric studies identified significant progressive reductions in frontal lobe gray matter as well as in frontal, temporal, and parietal lobe white matter^96^.

While the relationship between local brain age and more conventional structural neuroimaging features is a matter of further investigation, the above results suggest that local brain-predicted age may be sensitive to the underlying patterns of brain atrophy that are characteristic of various brain disorders. This implies that BrainAgeMap is a promising tool for the detailed delineation of advanced brain aging patterns through the direct quantification of region-wise deviations from the healthy brain aging trajectories. The resulting local brain age maps could be helpful in the recognition of various brain disorders. A recent meta-analysis showed that global brain-PAD is differentially expressed in SZ, bipolar disorder, and major depression^92^, and it has been suggested that global brain-PAD could aid the early differential diagnosis of SZ and bipolar disorder^6^. Our results, showing significantly advanced brain aging in circumscribed brain areas in schizophrenia, suggest that BrainAgeMap might further enhance the potential of brain age prediction to identify illness-specific brain structural alterations in neuropsychiatric disorders. Moreover, BrainAgeMap and similar models could provide novel insights into age-related atrophy patterns in brain diseases. In particular, local brain age prediction could be used to identify disease subtypes in a data-driven manner, by inputting local brain-PAD maps to clustering algorithms^4,37^. Data-driven methods applied to volumetric MRI data revealed different subgroups in various forms of dementia^97,98^. Incorporating information from local brain aging patterns might improve the effectiveness of these methods in disease subtyping. The observation that significantly greater accuracy can be achieved in predicting future conversion to AD using global brain age than hippocampal volumetric measures^99^ is indicative of the potential of regional brain age in disentangling the heterogeneity in neurodegenerative disorders.

When analyzing the local brain age maps output by a U-net model, a previous study has reported that the largest differences were observed in temporal lobe and subcortical regions when comparing AD patients with healthy controls, although significant group differences were found in most parts of the brain^37^. Using the same model, a later study demonstrated widespread advanced local brain aging in SZ that was most pronounced in frontal and temporal regions^100^. Our findings corroborate these results, although the group differences observed in our study appear to be more focal than previously reported. This is consistent with the recent successful application of an FCN model to generate precise, high resolution disease probability maps in AD, and highlights FCN’s potential to efficiently evaluate volumetric images in a contiguous manner without enforcing global structure onto local predictions^39^.

Besides the relatively small number of more recent investigations into local brain age prediction, several studies have attempted to draw conclusions regarding regional brain aging by identifying the brain features that affect the predictions of machine learning models trained for global brain age estimation. In the context of deep learning models using structural MRI scans in general, saliency methods^101^ are widely applied to highlight features in the scans that play an important role in model prediction^82,102–104^. These procedures provide an intuitive way of understanding model estimates through visual inspection^105^. However, some of these methods lack sensitivity to model parameters and the data generating process, as suggested by results showing that randomizing model parameters or data labels during training have little effect on the saliency maps^106^. In these cases, the methods are inadequate for explaining the relationship between the inputs and the outputs that the model learned, and while the maps may be visually compelling, they might expose the observer to the risk of confirmation bias^106^. Moreover, it is not clear whether non-salient regions in these maps are indeed not relevant for aging, or just too noisy or redundant with other structures^36^. In a recent study, Lee et al.^49^ developed a 3D DenseNet^107^ model for global brain age prediction based on T1-weighted images and performed a whole-brain voxel-wise regression analysis to investigate which brain regions’ alterations were associated with the global brain-PAD in different subject groups. Interestingly, in FTD, these associations were restricted to regions around the ventricles. This is in stark contrast with our current results showing significantly advanced local brain aging in FTD across the whole brain, with peaks in frontal and temporal regions. While there are several methodological differences between Lee et al.’s study and the current one, and a rigorous comparison of local brain age maps and visual explanations derived from global brain age models using various methods is outside the scope of the current study, these results, along with the above theoretical considerations raise the possibility that the two approaches might capture different aspects of the brain aging process.

In addition to cross-sectional comparisons of regional brain aging patterns between healthy subjects and participants with different brain disorders, we investigated longitudinal changes in local brain age in stable and progressive MCI. We observed significantly higher hippocampal brain-PAD in pMCI that was already present ∼2 years before conversion to AD, and found evidence for a steeper increase in brain-PAD across visits in the pMCI group. Our findings are in accordance with the results of longitudinal MRI morphometric studies showing faster brain atrophy in MCI-to-AD converters than in non-converters mainly in temporal lobe regions including the hippocampus^108–111^, and also in frontal cortical areas^111^, posterior cingulate, and precuneus^108^. A previous study demonstrated that individual differences in global brain-PAD reflect lifelong stable differences more than ongoing changes in brain structure in a sample devoid of individuals with MCI or AD.^112^ On the other hand, several investigations have provided evidence for longitudinal changes in global brain age in specific disease groups. Seminal studies revealed a significant increase at baseline and a greater accumulation of global brain-PAD over time in progressive compared to stable MCI^40,41^. This overall pattern of baseline and longitudinal differences in global brain-PAD has also been observed in SZ^113^ and type-2 diabetes mellitus^42^. To our knowledge, our study is the first to show accelerated local brain aging in progressive MCI subjects, thus demonstrating the feasibility of using locally estimated brain age to track neurodegenerative disease progression. This result, along with the observation of characteristic brain aging patterns in different disease groups, suggests that local brain age estimation might be useful in the diagnosis and monitoring of various brain disorders, particularly by aiding the identification of individuals at risk of faster brain structural deterioration and the evaluation of the effectiveness of therapeutic interventions.

We also examined the relationship between regional brain age and other physiological and cognitive measures to further examine the potential of local brain age as a putative aging biomarker. Our results revealed a significant positive correlation between local brain-PAD and tau uptake in the temporal lobe in AD subjects. In MCI, a tendency towards a similar effect was observed, whereas there was no significant association in the CN group. These results indicate that local brain age may be sensitive to abnormal metabolic function in neurodegenerative disease. Tau PET imaging has been shown to be effective in discriminating AD from other neurodegenerative disease states, and especially high diagnostic performance can be achieved when using a temporal meta-region of interest^46^. The accumulation of pathological tau proteins can be observed in several neurodegenerative disorders other than AD, and it has been assumed that abnormal tau spreading follows stereotypical spatiotemporal patterns in these diseases, although much uncertainty remains regarding the exact patterns^45^. Interestingly, a previous study observed significant positive associations between global brain-PAD derived from T1-weighted MRI scans and temporal lobe tau PET SUVR in MCI and AD, but not in frontotemporal dementia or dementia with Lewy bodies^49^. Given the potential of BrainAgeMap to identify distinctive patterns of advanced regional brain aging, future studies using this approach could investigate the putative associations between local brain age and tau uptake in different tauopathies.

Regarding the relationship between brain age and cognition, global brain-PAD has been shown to be associated with measures of general cognitive functioning in middle-aged and elderly adults^11,114,115^. With respect to neurodegeneration, significant correlations between global brain-PAD and neuropsychological test scores were also found in MCI and AD patients^41,82,116^. Here we attempted a detailed mapping of the associations between age-related structural changes and cognitive measures, finding a significant negative correlation in MCI between RAVLT Immediate—a proxy for verbal learning—and local brain-PAD mainly in medial temporal lobe regions and the fusiform gyrus. In AD subjects, the same pattern was evident across a more extensive set of brain regions. Prior evidence from machine-learning-based brain morphometric analysis also points to a direct link between neurodegenerative alterations in the medial temporal lobe and RAVLT performance. In particular, it has been shown that medial temporal lobe structures contributed most strongly to the predictions of an elastic net linear regression model trained to predict RAVLT Immediate scores from whole brain gray matter density maps in MCI and AD subjects and healthy controls^51^. Taken together, our findings indicate that differences in episodic memory function among people in a particular stage of neurocognitive impairment (MCI or AD) may be related to age-related neuroanatomical differences in regions including, but not limited to, medial temporal lobe structures. This suggests that BrainAgeMap and similar tools might be useful in characterizing individual structural alterations underlying neurocognitive impairment. In general, this approach may also provide new insights into the brain structural changes underlying cognitive dysfunction in dementia.

Taken together, our results suggest that BrainAgeMap is capable of detecting abnormal regional brain aging patterns in various disorders. Nevertheless, the current study has limitations, notably with respect to real-world application of BrainAgeMap. Our investigation, similar to most previous studies on brain age prediction using structural MRI data, relied on research-grade T1-weighted MRI scans obtained from open-access databases. To demonstrate the usefulness of using local brain age prediction in general and BrainAgeMap model in particular as a tool for screening, clinical decision making, and disease monitoring, it is essential to test such models on images from hospital databases which represent a greater variety of scanner types, imaging modalities (e.g., T2-weighted and diffusion weighted images), protocols, and patient populations^117^. Some recent investigations have demonstrated the feasibility of global brain age prediction using clinical-grade MRI scans^117–119^, but similar studies on local brain age prediction are yet to be conducted. BrainAgeMap has some desirable properties, such as its simple lightweight architecture which can be practical when model fine-tuning is required to accommodate other MRI sequences, and its capability to process images of arbitrary size. However, further testing on more heterogeneous datasets is required to examine its utility in medical applications. The frequent occurrence of image artifacts, especially as a result of head motion, is a common source of image quality degradation of routine clinical MRI examinations^120^. This is particularly problematic because head motion has a significant impact on global brain age estimation^57,121^. Here we used the normalized Euler number, a measure of image quality derived from Freesurfer, as covariate to control for this potential confound in the statistical analyses. However, further research is required to effectively assess and mitigate this problem in real-world settings. A further limitation of our study is that the training and validation sets consisted mostly of participants from Europe and North America, which might limit the generalizability of the findings to underrepresented populations. Global brain age prediction based on functional MRI and electroencephalography data has been shown to be sensitive to geographical and macrosocial factors^122^. In light of this finding, future research should investigate the role of these factors in regional brain aging by including more diverse samples.

## 5. Conclusions

In summary, we have introduced and validated a deep learning framework for mapping the topography of structural brain aging in detail. Our results demonstrate that these spatially-resolved aging patterns delineate neurobiologically meaningful, disease-specific pathways of neurodegeneration, linking macroscopic structural changes to underlying molecular pathology and cognitive decline. This approach may provide a powerful platform for stratifying patients in clinical trials, monitoring targeted therapeutic interventions, and ultimately advancing personalized neurology.

## Data and code availability statement

This study used data from publicly available datasets, some of which are available by application (see section S1.1. in Supplementary materials and methods for details). The model code will be made available upon the publication of the manuscript.

## Data Availability

This study used data from publicly available datasets, some of which are available by application (see section S1.1. in Supplementary materials and methods for details).

## Acknowledgements

This work was supported by the National Laboratory for Translational Neuroscience (TINL; project RRF-2.3.1-21-2022-00011; PI: Z.V.), the National Brain Research Program 3.0. by the Hungarian Academy of Sciences (NAP2022-I-1/2022; PI: Z.V.) and the Hungarian Research Network (HUN-REN; 298/4/2023/HF; PI: Z.V.). Data collection and sharing for this project was provided by the Cambridge Centre for Ageing and Neuroscience (CamCAN). CamCAN funding was provided by the UK Biotechnology and Biological Sciences Research Council (grant number BB/H008217/1), together with support from the UK Medical Research Council and University of Cambridge, UK. This research has been conducted using the UK Biobank Resource under Application Number 27236.

Data collection and sharing for this project was funded by the Alzheimer’s Disease Neuroimaging Initiative (ADNI) (National Institutes of Health Grant U01 AG024904) and DOD ADNI (Department of Defense award number W81XWH-12-2-0012). ADNI is funded by the National Institute on Aging, the National Institute of Biomedical Imaging and Bioengineering, and through generous contributions from the following: AbbVie, Alzheimer’s Association; Alzheimer’s Drug Discovery Foundation; Araclon Biotech; BioClinica, Inc.; Biogen; Bristol-Myers Squibb Company; CereSpir, Inc.; Cogstate; Eisai Inc.; Elan Pharmaceuticals, Inc.; Eli Lilly and Company; EuroImmun; F. Hoffmann-La Roche Ltd and its affiliated company Genentech, Inc.; Fujirebio; GE Healthcare; IXICO Ltd.; Janssen Alzheimer Immunotherapy Research & Development, LLC.; Johnson & Johnson Pharmaceutical Research & Development LLC.; Lumosity; Lundbeck; Merck & Co., Inc.; Meso Scale Diagnostics, LLC.; NeuroRx Research; Neurotrack Technologies; Novartis Pharmaceuticals Corporation; Pfizer Inc.; Piramal Imaging; Servier; Takeda Pharmaceutical Company; and Transition Therapeutics. The Canadian Institutes of Health Research is providing funds to support ADNI clinical sites in Canada. Private sector contributions are facilitated by the Foundation for the National Institutes of Health (www.fnih.org). The grantee organization is the Northern California Institute for Research and Education, and the study is coordinated by the Alzheimer’s Therapeutic Research Institute at the University of Southern California. ADNI data are disseminated by the Laboratory for Neuro Imaging at the University of Southern California.

Data were provided in part by OASIS-3: Longitudinal Multimodal Neuroimaging: Principal Investigators: T. Benzinger, D. Marcus, J. Morris; NIH P30 AG066444, P50 AG00561, P30 NS09857781, P01 AG026276, P01 AG003991, R01 AG043434, UL1 TR000448, R01 EB009352. AV-45 doses were provided by Avid Radiopharmaceuticals, a wholly owned subsidiary of Eli Lilly.

Data collection and sharing for this project was funded by the Frontotemporal Lobar Degeneration Neuroimaging Initiative (National Institutes of Health Grant R01 AG032306). The study is coordinated through the University of California, San Francisco, Memory and Aging Center. FTLDNI data are disseminated by the Laboratory for Neuro Imaging at the University of Southern California.

