## Supplementary Material for "Regional brain aging patterns reveal disease-specific pathways of neurodegeneration"

### 1. Supplementary materials and methods

#### 1.1. Details of the datasets

##### 1.1.1. Nathan Kline Institute-Rockland Sample (NKI-RS)

The Nathan Kline Institute-Rockland Sample (NKI-RS[1]) is a phenotypically rich dataset with state-of-the-art neuroimaging and genetic samples collected with the aim of delineating developmental trajectories across the human lifespan. The dataset consists of data obtained from individuals aged between 6 and 85 years. For the present study, we selected a single T1-weighted MRI scan from Session #1 for all subjects at or above 20 years of age. Image acquisition was performed on a 3T Siemens Magnetom TrioTim MRI scanner to acquire magnetization-prepared rapid gradient echo (MPRAGE) anatomical images (repetition time (TR) = 2500 ms, echo time (TE) = 3.5 ms, inversion time (TI) = 1200 ms, flip angle (FA) = 8°, field-of-view (FOV) = 256 × 256, slices = 190, voxel size = 1 mm isotropic) with or without 2-fold generalized autocalibrating partially parallel acquisitions (GRAPPA) acceleration. More details regarding the study can be found at: https://fcon_1000.projects.nitrc.org/indi/pro/nki.html.

##### 1.1.2. Information eXtraction from Images (IXI)

The Information eXtraction from Images (IXI) dataset contains structural MR images collected at three different hospitals in London, United Kingdom, from 590 normal, healthy subjects aged between 19 and 86 years. In the present study, we used T1-weighted scans collected at Hammersmith Hospital using a 3T Philips Medical Systems Intera scanner (Philips Medical Systems, Best, The Netherlands) from subjects at or above 20 years of age (TR = 9.6 ms, TE = 4.6 ms, FA = 8°, FOV = 256 × 256, slices = 150, voxel size = 0.9375 × 0.9375 × 1.2 mm). More details regarding the study can be found at: https://brain-development.org/ixi-dataset/.

##### 1.1.3. Southwest University Adult Lifespan Dataset (SALD)

The Southwest University Adult Lifespan Dataset (SALD) contains structural and resting-state functional MRI data, as well as basic phenotypic information, from 494 healthy adult participants aged between 19 and 80 years, collected with the aim of understanding age-related changes in brain structure and function[2]. Here we used the T1-weighted brain MRI scans of subjects at or above 20 years of age. These anatomical images were acquired on a 3T Siemens Trio MRI scanner (Siemens Medical, Erlangen, Germany) using an MPRAGE sequence (TR = 1900 ms, TE = 2.52 ms, TI = 900 ms, FA = 9°, FOV = 256 × 256, slices = 176, voxel size = 1 mm isotropic). More details can be found on the study's website at: https://fcon_1000.projects.nitrc.org/indi/retro/sald.html.

##### 1.1.4. Dallas Lifespan Brain Study (DLBS)

The Dallas Lifespan Brain Study (DLBS) is a major collection of anatomical MRI as well as PET scans along with cognitive, genetic, and demographic information from 315 participants aged between 18 and 89 years[3]. The data was collected with the purpose of understanding the antecedents of preservation and decline of cognitive functions. For the present study, we used the T1-weighted brain MRI scans of subjects at or above 20 years of age. Image acquisition was performed on a 3T Philips Achieva scanner using a T1-weighted 3D MPRAGE sequence (TR = 8.135 ms, TE = 3.7 ms, FA = 12°, FOV = 256 × 256, slices = 160, voxel size = 1 mm isotropic). More information regarding the dataset can be found on the study's website at: https://fcon_1000.projects.nitrc.org/indi/retro/dlbs.html.

##### 1.1.5. Cambridge Centre for Aging and Neuroscience (Cam-CAN)

The Cambridge Centre for Aging and Neuroscience (Cam-CAN) is a large-scale collaborative research project using epidemiological, cognitive, and neuroimaging data to understand the neurocognitive mechanisms underlying healthy cognitive ageing[4,5]. The data repository contains the structural brain MRI scans of 653 subjects aged between 18 and 88 years[5]. Quality control of the data was implemented on behalf of the Cam-CAN methods team by using semi-automated scripts. We used the T1-weighted brain MRI scans of subjects aged 20 years and above. The images were obtained on a 3T Siemens Magnetom TrioTim scanner using a 3D MPRAGE sequence (TR = 2250 ms, TE = 2.99 ms, TI = 900 ms, FA = 9°, FOV = 256 × 256, slices = 192, voxel size = 1 mm isotropic) and 2-fold GRAPPA acceleration. Further details about the Cam-CAN project can be found on the study's website: www.cam-can.org.

##### 1.1.6. UK Biobank (UKB)

UK Biobank (UKB) is a large population-based prospective study with over 500,000 participants recruited between 2006 and 2010 from across Great Britain. UKB comprises extensive phenotypic and genotypic characterization of participants using a great variety of methods including questionnaires, physical measurements, multimodal imaging, genome-wide genotyping and longitudinal follow-up of health-related outcomes[6]. A subset of the participants underwent MRI assessment starting from 2014. Data collection was performed on 3T Siemens Skyra MRI scanners (Siemens Healthineers, Erlangen, Germany), using a standard Siemens 32-channel RF receive head coil, at the UK Biobank imaging centers in Cheadle, Newcastle, and Reading[7]. 3D MPRAGE sequence (TR = 2000 ms, TE = 2.01 ms, TI = 880 ms, FA = 8°, FOV = 256 × 256, slices = 208, voxel size = 1 mm isotropic) with 2-fold GRAPPA acceleration was used for image acquisition. For the full description of the acquisition protocol, see:
https://biobank.ctsu.ox.ac.uk/crystal/refer.cgi?id=2367.
Image quality control on behalf of UK Biobank consisted of the rough manual review of T1 images supplemented by a beta-version automated quality control pipeline[7]. Subjects with an image that was deemed 'unusable' by the UK Biobank quality control pipeline were excluded from the present analyses. Moreover, subjects with certain ICD10 diagnoses (based on data field 41270) were excluded from the present study: Malignant neoplasms of meninges (ICD-10 code: C70) / brain (ICD-10 code: C71), Mental and behavioral disorders (ICD-10 codes: F00-F99), Diseases of the nervous system (ICD-10 codes: G00-G99), Symptoms and signs involving cognition, perception, emotional state and behaviour (ICD-10 codes: R40-46), Speech disturbances, not elsewhere classified (ICD-10 code: R47), Abnormal

findings on diagnostic imaging of CNS (ICD-10 code: R90), Intracranial injury (ICD-10 code: S06)[8]. In addition, we excluded subjects with a self-reported history of certain pathologies, namely: cancer (data field: 2453), stroke or heart attack (data field: 6150), blood clot in the lung or leg (data field: 6152), diabetes (data field: 2443), any long-standing illness, disability or infirmity (data field: 2188), or not having good or excellent self-reported health (data field: 2178-2.0)[9,10]. From the remaining pool of participants, 800 subjects were randomly selected using stratified sampling based on chronological age (data field: 21003-2.0) and sex (data field: 31-0.0).

###### 1.1.7. Alzheimer's Disease Neuroimaging Initiative (ADNI)

Data used in the preparation of this article were obtained from the Alzheimer's Disease Neuroimaging Initiative (ADNI) database (https://adni.loni.usc.edu). The ADNI was launched in 2003 as a public-private partnership, led by Principal Investigator Michael W. Weiner, MD. The primary goal of ADNI has been to test whether serial MRI, PET, other biological markers, and clinical and neuropsychological assessment can be combined to measure the progression of mild cognitive impairment (MCI) and early Alzheimer's disease (AD). For up-to-date information, see www.adni-info.org.

In the present study, we used structural MRI data from the ADNI-2 and ADNI-3 phases for model development and evaluation. In ADNI, subjects typically have several MRI scans as data collection took place over multiple sessions that may have spanned several years. MRI scans were acquired at multiple ADNI sites using various types of 3T MRI scanners from different vendors (General Electric (GE) Healthcare, Philips Medical Systems, Siemens Medical Solutions). We used the accelerated 3D T1-weighted MRI scans in the current study. A sagittal MPRAGE sequence was used in the case of Philips and Siemens scanners, whereas a sagittal inversion-recovery spoiled gradient recalled (IR-SPGR) sequence was used in the case of GE scanners. More details about the ADNI MRI scanner protocol are available online at: https://adni.loni.usc.edu/data-samples/adni-data/neuroimaging/mri/mri-scanner-protocols/. Each ADNI scan was assigned a diagnostic label of cognitively normal (CN), MCI (early and late MCI are not distinguished in the present study), or AD based on a diagnostic exam. As no diagnostic exam was performed at 3-month visits, these scans were included only if the subject had the same diagnosis at the screening and at the 6 month or 1-year visits. Additionally, MRI scans in the case of which more than 180 days passed between the acquisition date and the diagnostic exam date were removed (two 3-month CN scans in the case of which this interval was 181 days were kept in the analysis). Based on the available medical history, we excluded subjects with a history of seizures, abnormal electroencephalogram, brain tumor, ischemic stroke and intracranial hemorrhage, traumatic brain injury, brain aneurysm, arachnoid cyst, or Hashimoto encephalopathy. Subjects who had MCI or dementia due to etiology other than Alzheimer's Disease were not included in the current analyses.

##### 1.1.8. Open Access Series of Imaging Studies (OASIS)

The Open Access Series of Imaging Studies (OASIS) is a freely available multimodal dataset focusing on healthy aging and AD, available at: https://www.oasis-brains.org/. In the present study, we used data from the OASIS-3 data release. OASIS-3 includes structural and functional MRI, PET, neuropsychological and clinical data from 1098 participants, including cognitively normal individuals and individuals with early-stage AD, aged between 42 and 95 years[11]. All neuroimaging scans were conducted by the Knight Alzheimer Research Imaging Program at Washington University in St. Louis. MR imaging was performed on 3 different Siemens scanner models (Siemens Medical Solutions USA, Inc): Vision 1.5T, TIM Trio 3T (2 different scanners of this model), and BioGraph mMR PET-MR 3T. A 16-channel head coil was used on 1.5T scanners and a 20-channel head coil was used on 3T scanners with foam pad stabilizers placed next to the ears to reduce subject motion. Some participants had a vitamin-E fiducial marker placed on the left temple.

For the present analyses, we used T1-weighted images from the first visit from each subject with a corresponding clinical diagnostic entry available within one year. The matching of MRI visits and diagnostic entries were based on previous recommendations using The Extensible Neuroimaging Archive Toolkit (XNAT[12]) open source software available at http://github.com/NrgXnat/oasis-scripts. Subjects with a history of certain health issues registered up until the date of the MRI session -

stroke, seizures, traumatic brain injury, Parkinson's disease or other neurological or psychiatric condition were excluded from the analyses. In addition, participants with certain secondary diagnoses – active Parkinson's disease idiopathic, active or remote head trauma, active or remote seizure disorder – were excluded as well.

##### 1.1.9. Neuroimaging in Frontotemporal Dementia (NIFD)

NIFD is the nickname for the frontotemporal lobar degeneration neuroimaging initiative (FTLDNI, AG032306), which was funded by the National Institute of Aging and National Institute of Neurological Disorders and Stroke to characterize longitudinal clinical and imaging changes in frontotemporal lobar degeneration (FTLD). FTLDNI was funded through the National Institute of Aging, and started in 2010. The primary goals of FTLDNI were to identify neuroimaging modalities and methods of analysis for tracking FTLD and to assess the value of imaging versus other biomarkers in diagnostic roles. The Principal Investigator of NIFD was Dr. Howard Rosen, MD at the University of California, San Francisco. The data are the result of collaborative efforts at three sites in North America. For up-to-date information on participation and protocol, please visit http://memory.ucsf.edu/research/studies/nifd.

The NIFD dataset contains data from 345 subjects aged between 36 and 90 years. For the present study, we used the (non-accelerated) T1-weighted brain MRI scans of healthy control participants (CN) as well as individuals diagnosed with behavioral-variant frontotemporal dementia (bvFTD). Subjects with a diagnosis other than CN or bvFTD on any occasion were removed from the analysis. Image acquisition was performed on a 3T Siemens TrioTim scanner and 3T GE Discovery MR750 and Signa HDxt scanners.

##### 1.1.10. Center for Biomedical Research Excellence (COBRE)

The Center for Biomedical Research Excellence (COBRE) dataset contains structural MRI, resting-state functional MRI, and phenotypic data from 72 individuals diagnosed with schizophrenia (SZ) and 74 healthy control participants (CN), aged between 18 and 65 years. Participants were screened and excluded in case of a history of neurological disorder, history of mental retardation, history of severe head trauma with more than 5 minutes loss of consciousness, history of substance abuse or dependence within the last 12 months. Diagnostic information was collected using the Structured Clinical Interview used for DSM Disorders (SCID). More details about the dataset are available online at: https://fcon_1000.projects.nitrc.org/indi/retro/cobre.html.

For the present study, we selected the T1-weighted MRI scans of subjects at or above 20 years of age. Image collection was performed on a Siemens TrioTim +T scanner using a multi-echo MPRAGE sequence (TR = 2530 ms, TE = 1.64, 3.5, 5.36, 7.22, 9.08 ms, TI = 900 ms, FA = 7°, FOV = 256 × 256, slices = 192, voxel size = 1 mm isotropic).

#### 1.2. Demographic characteristics

Table S1. Demographic characteristics of the subjects from different datasets included in the training, validation, and test sets of the current study. Note that the Hippocampal Brain Age dataset and the Tau PET dataset contain subject data from the Alzheimer’s Disease Neuroimaging Initiative (ADNI) database, from the ADNI-2 and ADNI-3 phases. AD: Alzheimer’s Disease; bvFTD: behavioral-variant frontotemporal dementia; Cam-CAN: Cambridge Centre for Aging and Neuroscience; CN: cognitively normal / healthy control subjects; COBRE: Center for Biomedical Research Excellence; DLBS: Dallas Lifespan Brain Study; IXI: Information eXtraction from Images; MCI: mild cognitive impairment; NIFD: Neuroimaging in Frontotemporal Dementia; NKI-RS: Nathan Kline Institute-Rockland Sample; OASIS: Open Access Series of Imaging Studies; pMCI: progressive mild cognitive impairment; RAVLT: Rey’s Auditory Verbal Learning Test; SALD: Southwest University Adult Lifespan Dataset; sMCI: stable mild cognitive impairment; SZ: schizophrenia; UKB: UK Biobank.

| Dataset | Subset | Subject Group | Number of images | Number of subjects (females) | Age range (years) | Mean age ± standard deviation (years) |
|---|---|---|---|---|---|---|
| NKI-RS | Train | All | 133 | 133 (52) | 20 - 83 | 41.98 ± 17.30 |
| | Valid | All | 15 | 15 (6) | 20 - 70 | 39.33 ± 15.39 |
| IXI | Train | All | 160 | 160 (82) | 20 - 82 | 47.24 ± 16.90 |
| | Valid | All | 18 | 18 (9) | 24 - 72 | 47.78 ± 15.50 |
| SALD | Train | All | 437 | 437 (272) | 20 - 80 | 45.49 ± 17.29 |
| | Valid | All | 49 | 49 (31) | 20 - 77 | 46.31 ± 17.27 |
| DLBS | Train | All | 275 | 275 (170) | 20 - 89 | 53.49 ± 19.84 |
| | Valid | All | 31 | 31 (20) | 22 - 86 | 52.58 ± 19.92 |
| Cam-CAN | Train | All | 563 | 563 (289) | 20 - 88 | 54.39 ± 18.07 |
| | Valid | All | 63 | 63 (33) | 24 - 87 | 53.68 ± 17.87 |
| UKB | Train | All | 705 | 705 (398) | 46 - 81 | 63.19 ± 7.53 |
| | Valid | All | 79 | 79 (45) | 49 - 79 | 63.67 ± 7.81 |
| ADNI-2 | Train | CN | 665 | 152 (83) | 58 - 94 | 74.56 ± 6.40 |
| | Valid | CN | 86 | 20 (11) | 64 - 88 | 74.31 ± 6.46 |
| | Test | CN | 100 | 20 (11) | 64 - 88 | 74.45 ± 6.44 |
| OASIS-3 | Test | CN | 55 | 55 (29) | 60 - 91 | 76.04 ± 7.55 |
| | | MCI | 55 | 55 (29) | 61 - 90 | 75.80 ± 6.93 |
| | | AD | 55 | 55 (29) | 59 - 96 | 76.55 ± 8.78 |
| NIFD | Test | CN | 73 | 73 (23) | 39 - 76 | 62.64 ± 7.04 |
| | | bvFTD | 73 | 73 (23) | 46 - 76 | 62.35 ± 6.35 |
| COBRE | Test | CN | 65 | 65 (15) | 20 - 65 | 36.72 ± 11.28 |
| | | SZ | 65 | 65 (13) | 20 - 65 | 39.38 ± 13.19 |
| Hippocampal Brain Age | Test | sMCI | 93 | 31 (13) | 59 - 90 | 73.64 ± 6.97 |
| | | pMCI | 93 | 31 (13) | 58 - 90 | 73.89 ± 8.01 |
| Tau PET | Test | CN | 34 | 34 (12) | 62 - 90 | 77.79 ± 6.82 |
| | | MCI | 34 | 34 (12) | 61 - 93 | 77.59 ± 6.95 |

| | | | | | | |
|---|---|---|---|---|---|---|
| | | AD | 34 | 34 (12) | 57 - 94 | 78.56 ± 8.43 |
| RAVLT | Test | CN | 188 | 188 (85) | 58 - 91 | 74.70 ± 6.90 |
| | | MCI | 188 | 188 (78) | 56 - 94 | 75.55 ± 7.90 |
| | | AD | 188 | 188 (78) | 56 - 94 | 75.53 ± 7.93 |

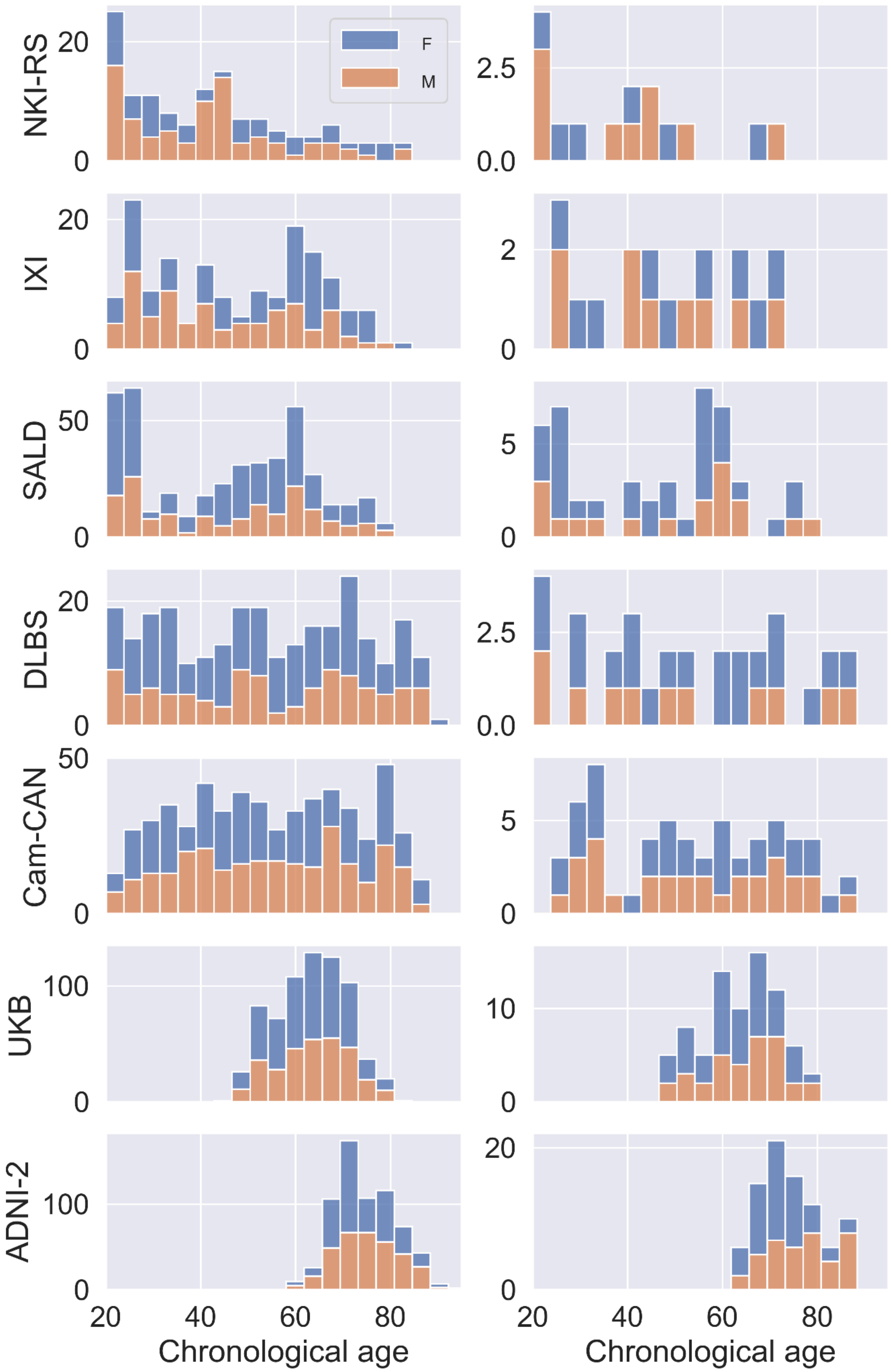


Fig. S1. Distribution of chronological age (years) and sex (F: female, M: male) of subjects from different datasets included in the training (left column) and validation sets (right column). Image count is plotted on the y axis for each dataset. ADNI: Alzheimer's Disease Neuroimaging Initiative; Cam-CAN: Cambridge Centre for Aging and Neuroscience; DLBS: Dallas Lifespan Brain Study; IXI: Information eXtraction from Images; NKI-RS: Nathan Kline Institute-Rockland Sample; SALD: Southwest University Adult Lifespan Dataset; UKB: UK Biobank.

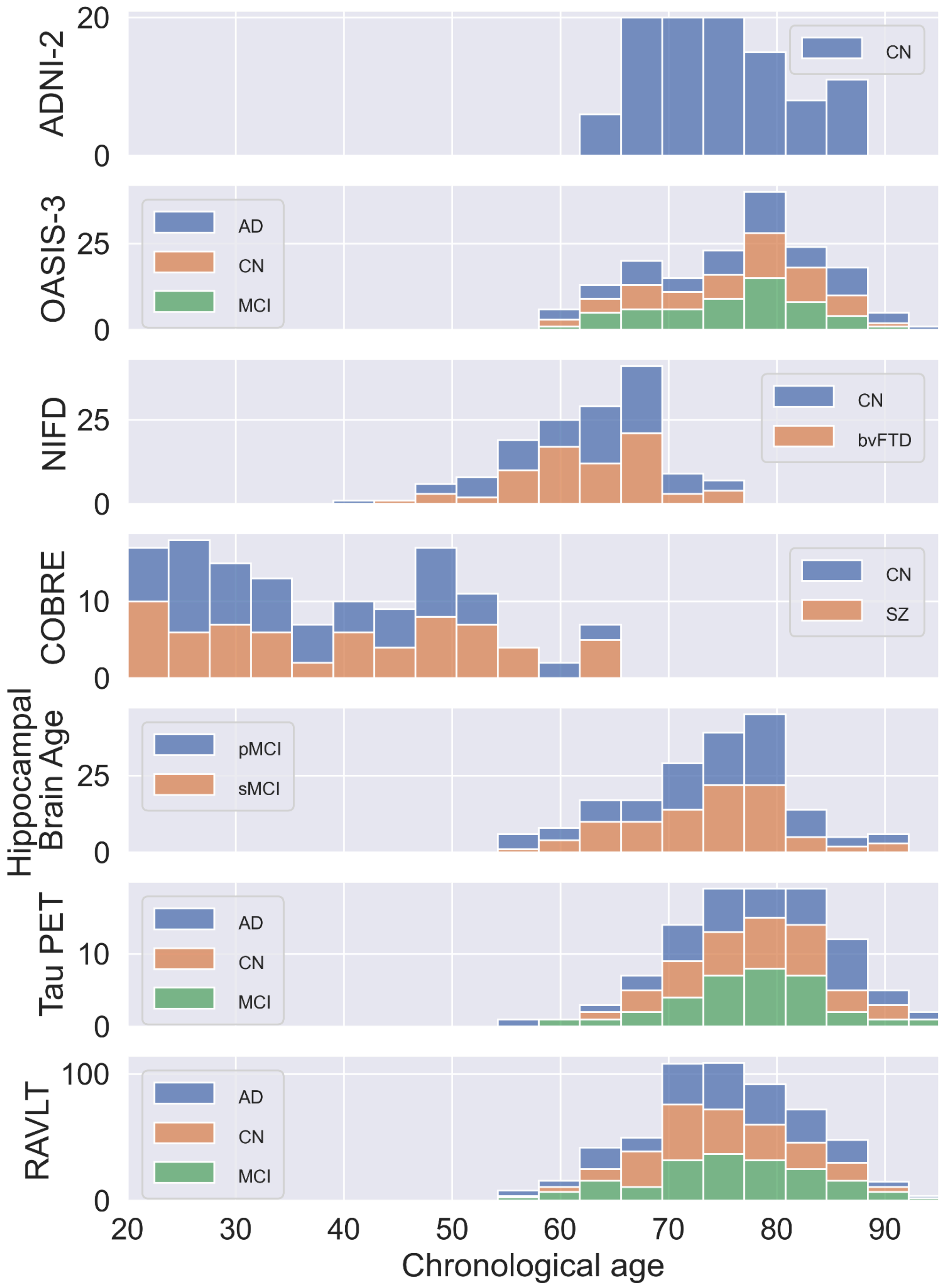


Fig. S2. Distribution of chronological age in the different test datasets and subject groups. Image count is plotted on the y axis for each dataset. AD: Alzheimer's disease; ADNI: Alzheimer's Disease Neuroimaging Initiative; bvFTD: behavioral-variant frontotemporal dementia; CN: cognitively normal / healthy control subjects; COBRE: Center for Biomedical Research Excellence; MCI: mild cognitive impairment; NIFD: Neuroimaging in Frontotemporal Dementia; OASIS: Open Access Series of Imaging Studies; pMCI: progressive mild cognitive impairment; RAVLT: Rey's Auditory Verbal Learning Test; sMCI: stable mild cognitive impairment; SZ: schizophrenia.

##### 1.3. Short description of the Rey's Auditory Verbal Learning Test (RAVLT) and the distribution of test scores

In RAVLT[13], a list of 15 words (list A) is read to the participant who is then immediately asked to recall as many words as possible. This procedure is repeated five times (Trials 1-5), after which a new list of 15 words (list B) is read to the participant, followed by an immediate instruction to recall list B. Then, the participant is asked to recall the words from list A (Trial 6). After a delay of 30 minutes, the participant is asked again to recall the words from list A.

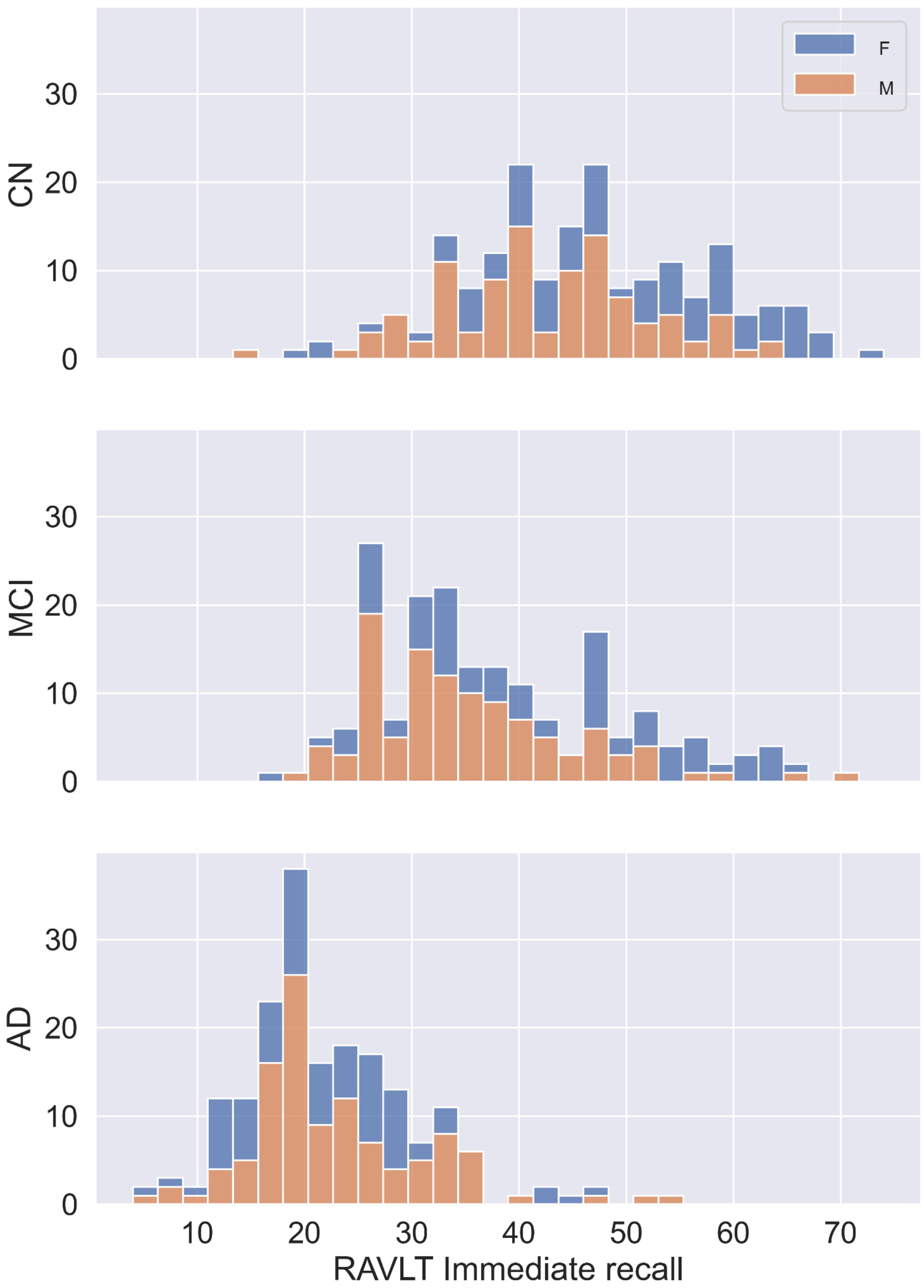


Fig. S3. Distribution of the Immediate summary score in Rey's Auditory Verbal Learning Test (RAVLT) in cognitively normal (CN) subjects and subjects with mild cognitive impairment (MCI) and Alzheimer's disease (AD). F: female; M: male.

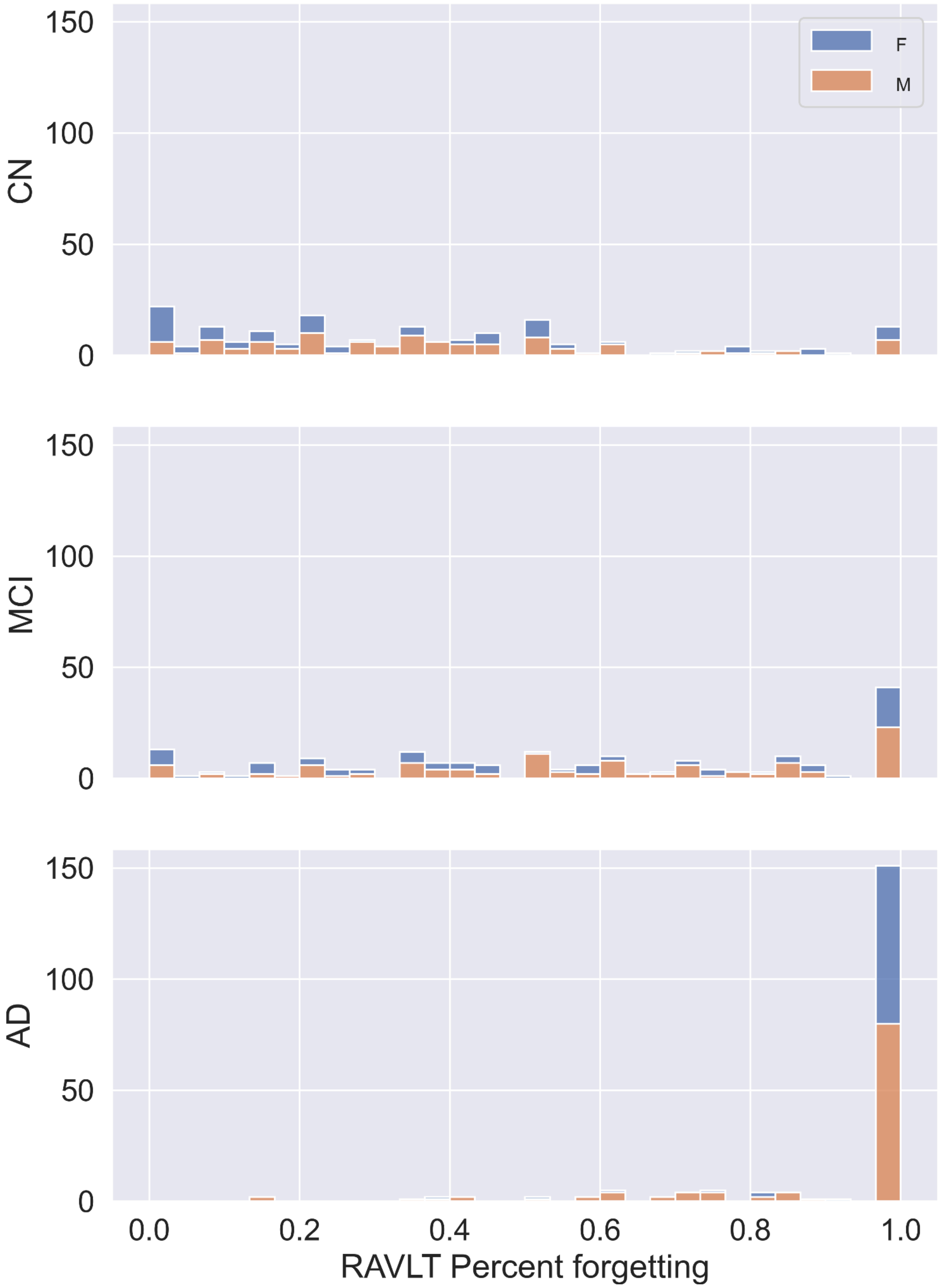


Fig. S4. Distribution of the Percent Forgetting summary score in Rey's Auditory Verbal Learning Test (RAVLT) in cognitively normal (CN) subjects and subjects with mild cognitive impairment (MCI) and Alzheimer's disease (AD). F: female; M: male.

#### 1.4. Details of the neural network architectures

Table S2. Architecture of the BrainAgeMap base neural network model used for local (regional) brain age prediction. 3D convolutional layers are specified using the format 'CONV3D(I×J×K@F)', where I, J, and K denote the depth, height, and width of the 3D convolution window, respectively, and F denotes the number of filters. Similarly, the I, J, and K in 'MAXPOOL(I×J×K)' denote the dimensions of the max pooling window. The flattening and dense output layers marked with a star (*) were included in the model (3D-CNN) only during model training (using cuboid patches as input). The convolutional output layer marked with a dagger (†) was included in the model (3D-FCN) only during inference (using padded whole images as input). The convolutional output layer (conv_output) used during inference has the same weights as the ones learned by the dense output layer (dense_output) during training. ReLU: rectified linear unit.

| Layer name | Layer type | Stride | Output shape (patch) | Output shape (whole image) |
|---|---|---|---|---|
| input | Input | - | 47×47×47×1 | 203×235×202×1 |
| conv_1 | CONV3D(4×4×4@20) | 1 | 44×44×44×20 | 200×232×199×20 |
| max_pool_1 | MAXPOOL(2×2×2) | 1 | 43×43×43×20 | 199×231×198×20 |
| bn_1 | Batch normalization | - | 43×43×43×20 | 199×231×198×20 |
| leaky_relu_1 | Activation (leaky ReLU) | - | 43×43×43×20 | 199×231×198×20 |
| dropout_1 | Dropout (rate = 0.1) | - | 43×43×43×20 | 199×231×198×20 |
| conv_2 | CONV3D(4×4×4@40) | 1 | 40×40×40×40 | 196×228×195×40 |
| max_pool_2 | MAXPOOL(2×2×2) | 2 | 20×20×20×40 | 98×114×97×40 |
| bn_2 | Batch normalization | - | 20×20×20×40 | 98×114×97×40 |
| leaky_relu_2 | Activation (leaky ReLU) | - | 20×20×20×40 | 98×114×97×40 |

| | | | | |
|---|---|---|---|---|
| dropout_2 | Dropout (rate = 0.1) | - | 20×20×20×40 | 98×114×97×40 |
| conv_3 | CONV3D(3×3×3@80) | 1 | 18×18×18×80 | 96×112×95×80 |
| max_pool_3 | MAXPOOL(2×2×2) | 2 | 9×9×9×80 | 48×56×47×80 |
| bn_3 | Batch normalization | - | 9×9×9×80 | 48×56×47×80 |
| leaky_relu_3 | Activation (leaky ReLU) | - | 9×9×9×80 | 48×56×47×80 |
| dropout_3 | Dropout (rate = 0.1) | - | 9×9×9×80 | 48×56×47×80 |
| conv_4 | CONV3D(3×3×3@160) | 1 | 7×7×7×160 | 46×54×45×160 |
| max_pool_4 | MAXPOOL(2×2×2) | 1 | 6×6×6×160 | 45×53×44×160 |
| bn_4 | Batch normalization | - | 6×6×6×160 | 45×53×44×160 |
| leaky_relu_4 | Activation (leaky ReLU) | - | 6×6×6×160 | 45×53×44×160 |
| dropout_4 | Dropout (rate = 0.5) | - | 6×6×6×160 | 45×53×44×160 |
| conv_5 | CONV3D(6×6×6@30) | 1 | 1×1×1×30 | 40×48×39×30 |
| leaky_relu_5 | Activation (leaky ReLU) | - | 1×1×1×30 | 40×48×39×30 |
| dropout_5 | Dropout (rate = 0.5) | - | 1×1×1×30 | 40×48×39×30 |
| *flatten | Flattening | - | 30 | - |
| *dense_output | Fully connected (Dense) | - | 1 | - |
| †conv_output | CONV3D(1×1×1@1) | 1 | - | 40×48×39×1 |

Table S3. Architecture of the 3D convolutional neural network (3D-CNN) used as a BrainAgeMap head model variant for global brain age prediction. 3D convolutional layers are specified using the format ‘CONV3D(I×J×K@F)’, where I, J, and K denote the depth, height, and width of the 3D convolution window, respectively, and F denotes the number of filters. Similarly, the I, J, and K in ‘MAXPOOL(I×J×K)’ denote the dimensions of the max pooling window. ReLU: rectified linear unit.

| Layer name | Layer type | Stride | Output shape |
|---|---|---|---|
| input | Input | - | 40×48×39×1 |
| conv_1 | CONV3D(3×3×3@16) | 1 | 40×48×39×16 |
| bn_1 | Batch normalization | - | 40×48×39×16 |
| leaky_relu_1 | Activation (leaky ReLU) | - | 40×48×39×16 |
| max_pool_1 | MAXPOOL(2×2×2) | 2 | 20×24×19×16 |
| conv_2 | CONV3D(3×3×3@32) | 1 | 20×24×19×32 |
| bn_2 | Batch normalization | - | 20×24×19×32 |
| leaky_relu_2 | Activation (leaky ReLU) | - | 20×24×19×32 |
| max_pool_2 | MAXPOOL(2×2×2) | 2 | 10×12×9×32 |
| conv_3 | CONV3D(3×3×3@32) | 1 | 10×12×9×32 |
| bn_3 | Batch normalization | - | 10×12×9×32 |
| leaky_relu_3 | Activation (leaky ReLU) | - | 10×12×9×32 |
| max_pool_3 | MAXPOOL(2×2×2) | 2 | 5×6×4×32 |
| conv_4 | CONV3D(3×3×3@64) | 1 | 5×6×4×64 |
| bn_4 | Batch normalization | - | 5×6×4×64 |
| leaky_relu_4 | Activation (leaky ReLU) | - | 5×6×4×64 |

| | | | |
|---|---|---|---|
| gap | Global average pooling | - | 64 |
| dropout | Dropout (rate = 0.5) | - | 64 |
| dense_output | Fully connected (Dense) | - | 1 |

### 2. Supplementary results

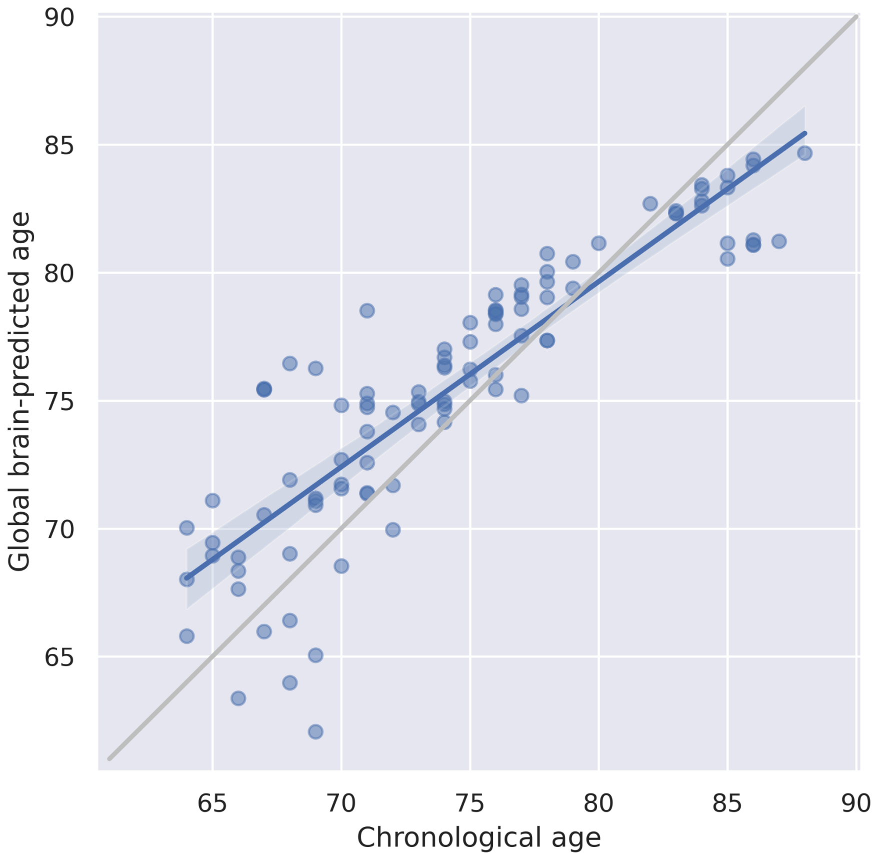


Fig. S5. Correlation between chronological age and global brain-predicted age (years) in cognitively normal (CN) subjects in the ADNI-2 test set. Blue and gray lines correspond to the regression lines and the lines of identity, respectively. The shaded areas represent the 95% confidence intervals.

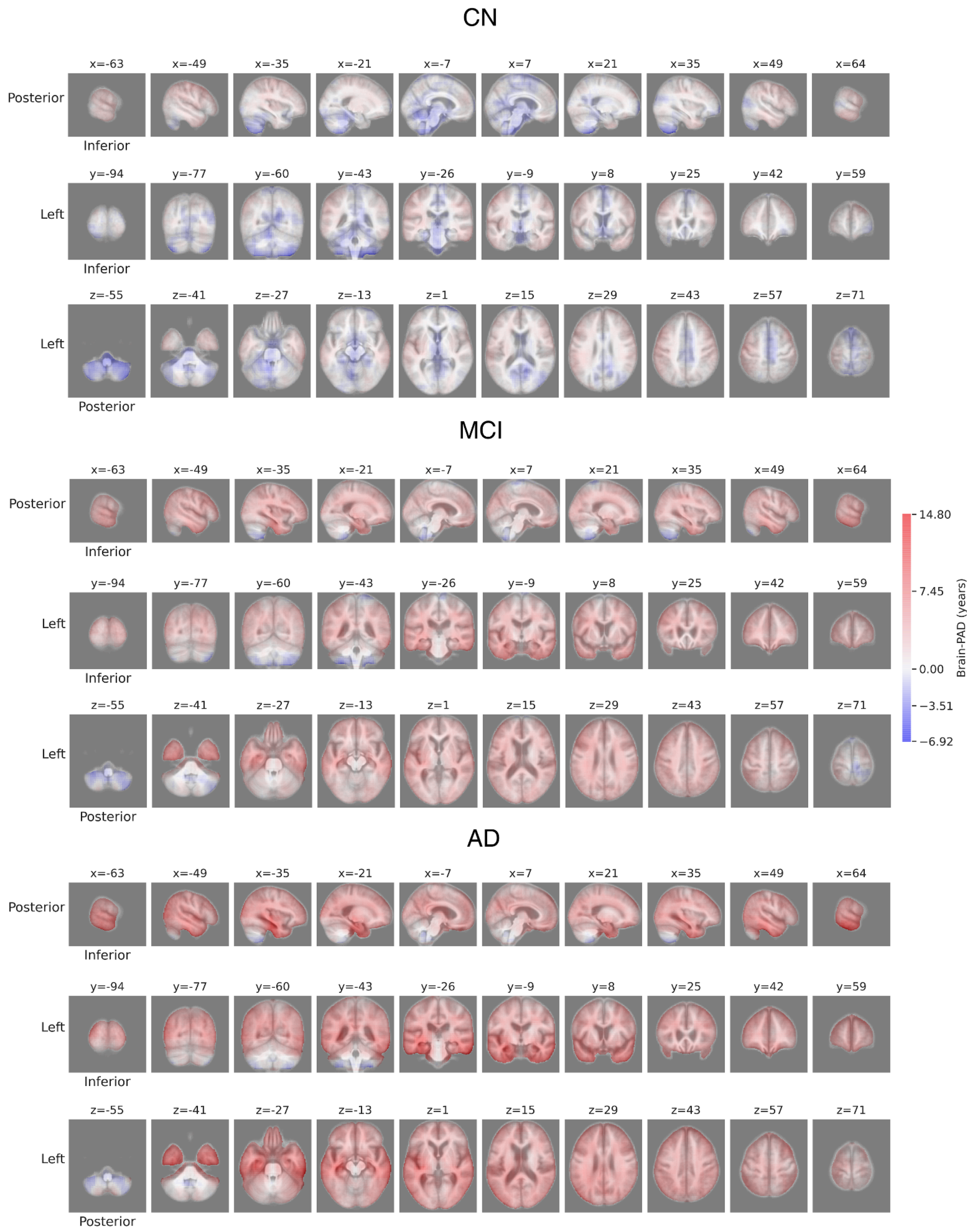


Fig. S6. Grand average local brain-PAD maps in the OASIS-3 dataset. Brain-PAD (brain-predicted minus chronological age) was calculated for each voxel and averaged across subjects in the cognitively normal (CN), mild cognitive impairment (MCI) and Alzheimer's disease (AD) groups.

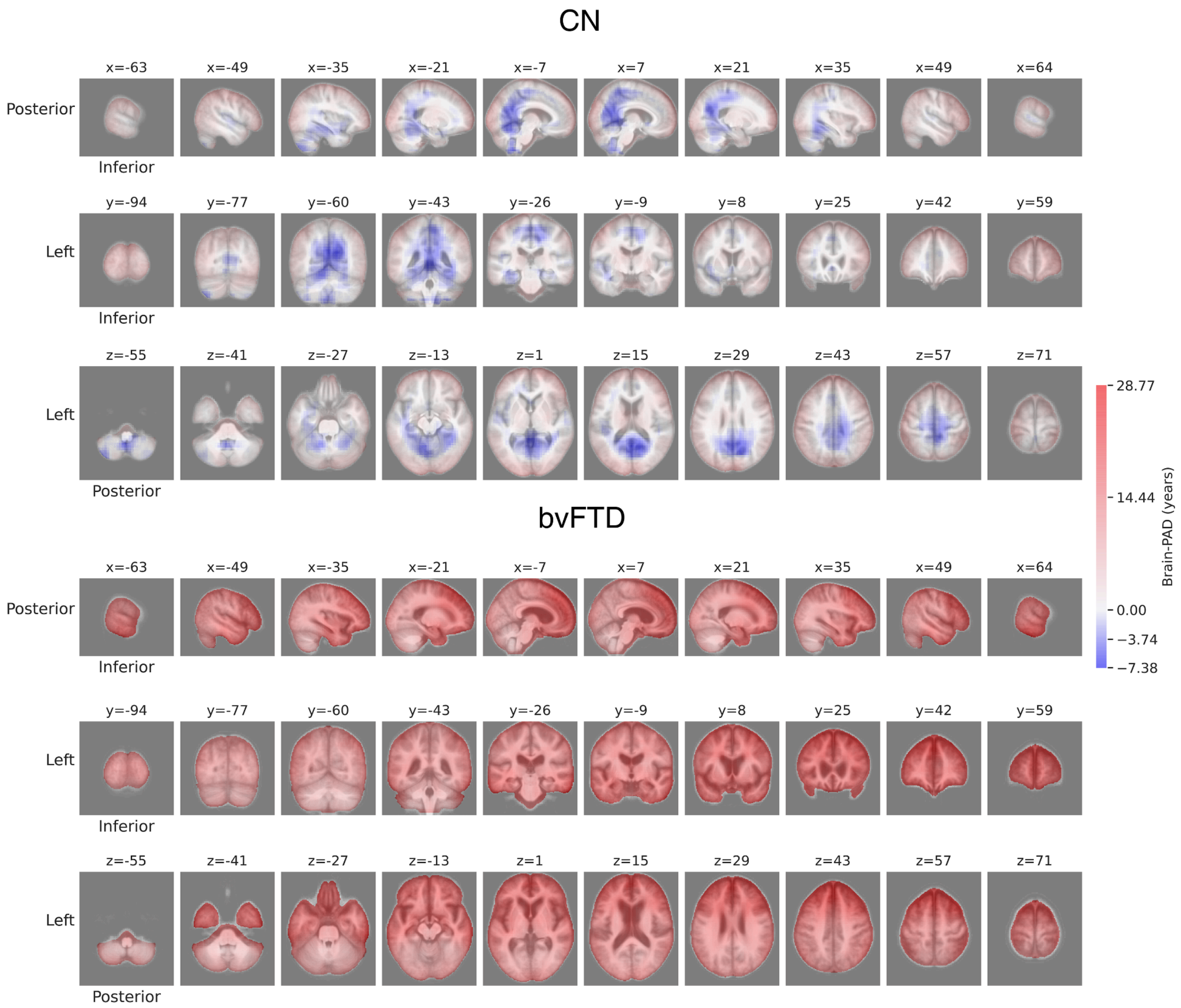


Fig. S7. Grand average local brain-PAD maps in the NIFD dataset. Brain-PAD (brain-predicted minus chronological age) was calculated for each voxel and averaged across subjects in the cognitively normal (CN) and behavioral-variant frontotemporal dementia (bvFTD) groups.

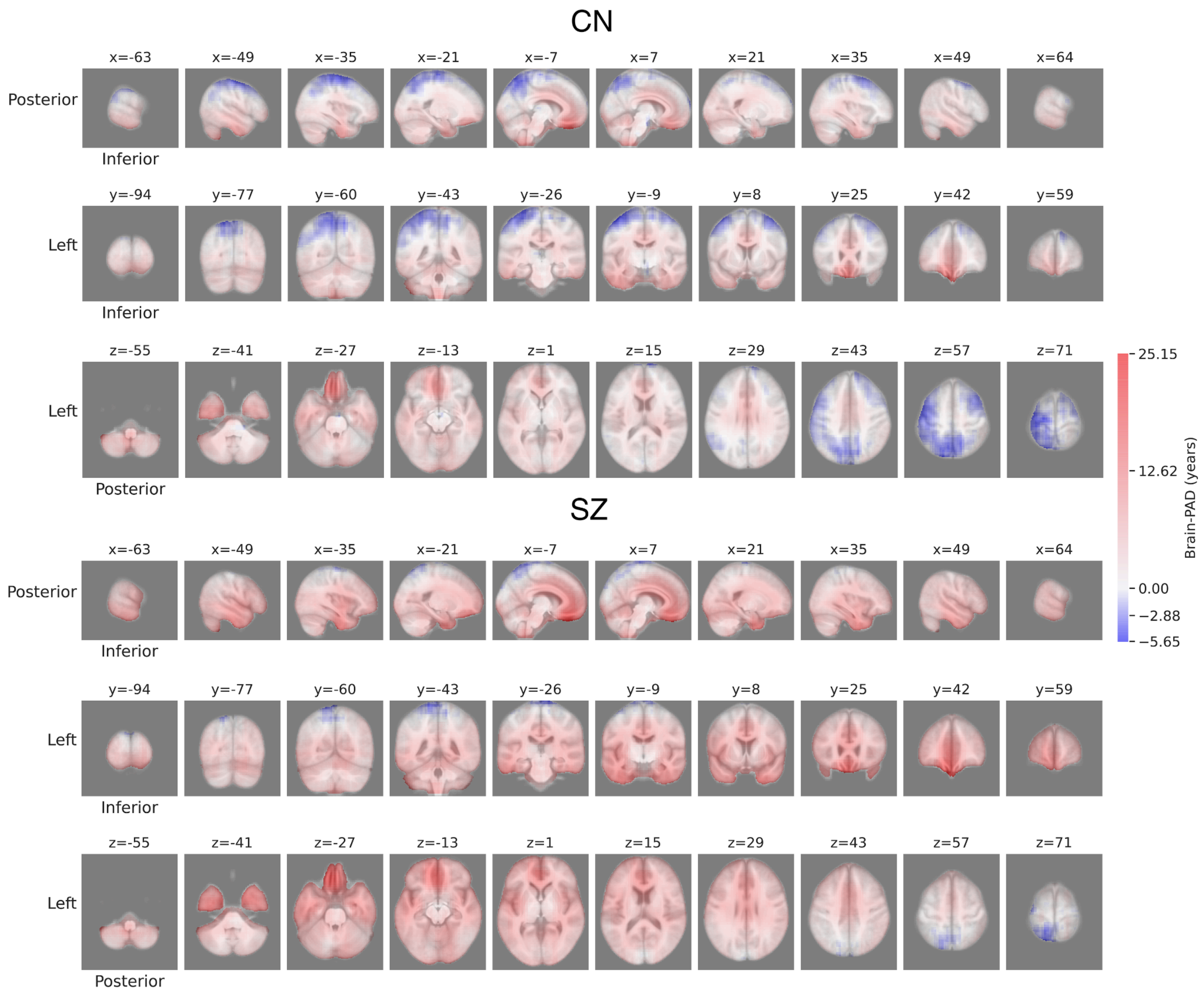


Fig. S8. Grand average local brain-PAD maps in the COBRE dataset. Brain-PAD (brain-predicted minus chronological age) was calculated for each voxel and averaged across subjects in the cognitively normal (CN) and schizophrenia (SZ) groups.

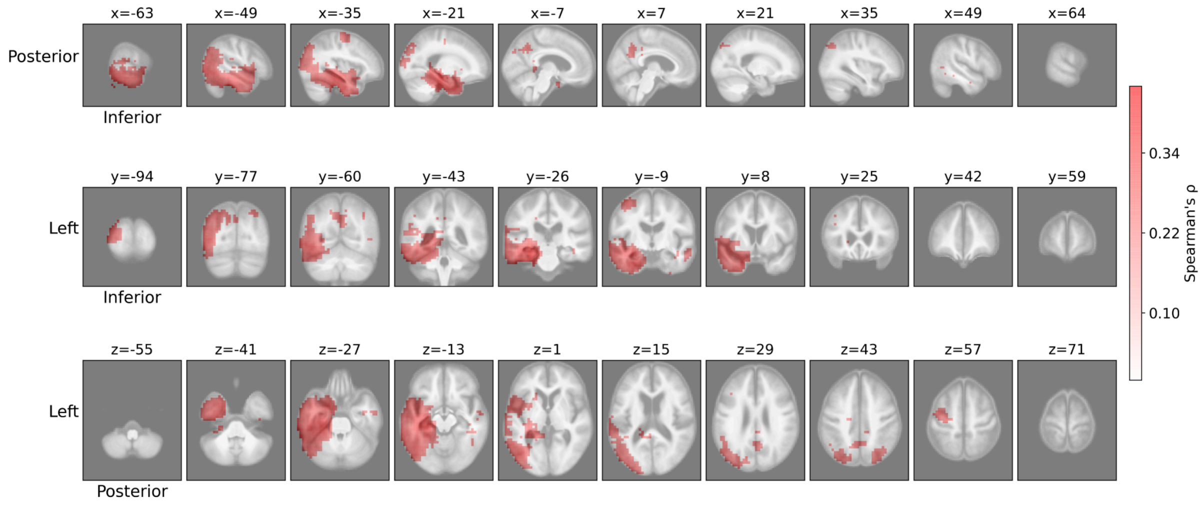


Fig. S9. Topographical distribution of the correlation coefficient (Spearman's ρ) when assessing the correlation between local brain-PAD (brain-predicted minus chronological age) and Percent Forgetting in Rey's Auditory Verbal Learning Test (RAVLT) in subjects with Alzheimer's disease (AD). The coefficient is shown only for voxels where the correlation was significant after correction for multiple comparisons.

Table S4. Linear mixed-effects model predicting local brain-PAD.

**Fixed effects**

| **Predictor** | **Estimate** | **SE** | **df** | **t** | **p** | **CI** |
|---|---|---|---|---|---|---|
| Intercept | 25.72 | 5.77 | 60.74 | 4.46 | < .001 | [13.30,38.21] |
| Visit number (absolute) | 0.06 | 0.23 | 78.26 | 0.28 | .778 | [-0.38,0.51] |
| Group (Progressive vs. Stable) | 7.21 | 1.39 | 53.76 | 5.21 | < .001 | [4.52,9.91] |
| Euler number (normalized) | -0.10 | 0.40 | 129.43 | -0.25 | .805 | [-0.89,0.70] |
| Age | -0.32 | 0.08 | 63.84 | -4.02 | < .001 | [-0.50,-0.15] |
| Visit × Group interaction | 1.00 | 0.30 | 59.92 | 3.38 | .001 | [0.42,1.58] |

**Random effects**

| **Group** | **Effect** | **Variance** | **SD** | **Corr** |
|---|---|---|---|---|
| Subject_ID | Intercept | 26.92 | 5.19 | |
| | Visit number | 0.77 | 0.88 | -0.45 |
| Residual | | 1.20 | 1.10 | |

Table S5. Linear mixed-effects model predicting global brain-PAD.

**Fixed effects**

| **Predictor** | **Estimate** | **SE** | **df** | **t** | **p** | **CI** |
|---|---|---|---|---|---|---|
| Intercept | 43.03 | 4.36 | 60.90 | 9.88 | < .001 | [33.51,52.63] |
| Visit number (absolute) | 0.13 | 0.15 | 83.84 | 0.91 | .364 | [-0.16,0.42] |
| Group (Progressive vs. Stable) | 3.73 | 1.09 | 52.83 | 3.42 | .001 | [1.61,5.86] |
| Euler number (normalized) | -0.12 | 0.28 | 134.83 | -0.44 | .663 | [-0.67,0.45] |
| Age | -0.59 | 0.06 | 63.72 | -9.68 | < .001 | [-0.72,-0.46] |
| Visit × Group interaction | 0.45 | 0.19 | 58.70 | 2.41 | .019 | [0.09,0.81] |

**Random effects**

| **Group** | **Effect** | **Variance** | **SD** | **Corr** |
|---|---|---|---|---|
| Subject_ID | Intercept | 16.87 | 4.11 | |
| | Visit number | 0.19 | 0.44 | -0.61 |
| Residual | | 0.68 | 0.82 | |
